## Supplementary Information for "Genome-wide association analysis and Mendelian randomization proteomics identify drug targets for heart failure"

### SUPPLEMENTARY METHODS

#### Genotyping, Quality Control, and Imputation of Genetic Data

For the data obtained from the Million Veteran Program (MVP), DNA was extracted from participants' blood and genotyped using the MVP 1.0 Genotyping Array, a customized Affymetrix Axiom biobank array. The MVP 1.0 Genotyping Array is enriched for both common and rare genetic variants of clinical significance. The Axiom genotyping platform uses a two-color, litigation assay with 30-mer oligonucleotide probes. The probes are synthesized *in situ* onto the surface of a microarray substrate. The design features of the MVP 1.0 Genotyping Array and additional details of the Axiom Genotyping Platform have been previously described[1]. Genotype calling was performed on unprocessed Axiom genotype data for 485,856 unique samples, followed by normalization for mitigating plate-to-plate variation[1]. As part of the quality control procedure, samples that were duplicated, with heterozygosity greater than expected, with greater than 2.5% missing genotype calls, or with discordance between genetically inferred sex and phenotypic gender were excluded from further analyses [1]. Samples were checked for contamination[1]. Excess relatedness was assessed using the relatedness inference software KING[2], and samples that did not meet the threshold for relatedness were removed from further analysis, as well as variants that exhibited genotype missingness greater than 5% or those that deviated from their expected allele frequency as observed in the 1000 Genomes reference data. A full detailed summary of the quality control procedure for the MVP 1.0 Array can be found in the Supplementary Materials of the publication[1]. Pre-phasing was executed using the EAGLE Version 2.4 software[3], and imputation was executed using the 1000 Genomes Phase 3 reference panel using Minimac4[4]. Imputation performance was assessed, and variants that had poor quality as determined by $r^2 < 0.3$ were removed from further analyses.

All studies included in the HERMES Consortium utilized high-density genotyping arrays. A detailed table summarizing the genotyping, quality control, imputation, and analysis across the 29 distinct datasets included in the HERMES Consortium has been previously described[5]. For quality control, the per variant call rate and the per sample call rate across all studies was at least greater than 90[5]. The MAF threshold ranged from >0% to 1% across studies[5]. Imputation was performed using at least one of the following reference panels: 1000 Genomes (Phase 1 or Phase 3), Hapmap 2 NCBI build 36, Haplotype Reference Consortium, the Estonian Whole-Genome Sequence reference, or a reference of 15,220 whole genomes sequenced by Icelanders (deCODE). Phasing was conducted using Eagle[3], MaCH[6], and SHAPEIT[7] software, and imputation was conducted using mimimac2[8] and IMPUTE2[9]. Imputation details varied by study, and have been previously documented[5].

#### Phenotyping of heart failure

Across all 26 cohorts of the HERMES Consortium, cases with HF were identified by a clinical diagnosis of HF of any etiology, as determined by physician diagnosis or adjudication, ICD codes, and imaging, and controls were participants without a clinical diagnosis of HF. There was no inclusion criteria based on LV ejection fraction. More descriptive definitions for heart failure as well as inclusion criteria by study included within the HERMES Consortium has been previously documented[5]. In the MVP, HF patients were identified as those with an International Classification of Diseases (ICD)-9 code of 428.x or ICD-10 code of I50.x *and* an echocardiogram performed within 6 months of diagnosis (median time period from diagnosis to echocardiography was 3 days, interquartile range 0-32 days). In the MVP, comorbid conditions were curated using ICD-9 or ICD-

10 codes.

**Genebass-UK Biobank**

Genebass is a publicly available resource of exome-based association statistics derived from the UK Biobank study. The dataset encompasses 4,529 phenotypes with gene-based and single-variant testing across 426,370 individuals with exome sequence data from the UK Biobank. The burden tests available in Genebass were performed using the mixed model framework implemented in SAIGE-GENE, which includes single-variant tests and gene-based burden, SKAT, and SKAT-O tests. We used the results from the gene-based burden analysis. These results were derived using SAIGE-GENE and fitting a model adjusted by the fixed factors age, sex, $age^2$, the first 20 principal components, and the interaction terms age*sex and $age^2$*sex.

**Epigraph Strategy**

In order to investigate the current epidemiological knowledge about the biomedical functions of the hit genes in association with HF, we used the EpiGraphDB database[11]. From the variety of API that are provided on the database, we used the *query_epigraphdb* to query the biomedical and epidemiological relationships curated in the database. On July 8, 2022, we queried the database to identify associations between the hit genes and a variety of medical endpoints including cardiovascular related outcomes and cardiovascular disease risk factors. These endpoints include: Apolipoproteins, Arrhythmias, Atrial fibrillation, BMI, C-reactive protein, Cardiac, Cardiac allograft, Cardiac differentiation, cardiac fibroblast, Cardiac hypertrophy, Cardiac injury, Cardiomyocyte, Cardiomyopathy, cardiopulmonary, Cardiovascular, Cholesterol, Congenital Cardiomyopathy, Congestive heart failure, congestive heart failure, coronary heart disease, CRP, CVD, Cytokines, Diabetes, Diastolic, Dyslipidemia, Ejection Fraction, Endothelial, GRAFT, Heart, Heart attack, Heart Disorders, Heart failure, HF, HFpEF, HFrEF, High-risk hypertension, Hypercholesterolemia, Hypertension, Hypertrophic cardiomyopathy, Immunosuppression, Inflammation, Kidney disease, Kidney failure, Lipodystrophy, LV ejection, Mitochondrial, Myocardial disease, Myocardial infarction, Myocardium, Obesity, Statin, Stroke, Systolic, Triglycerides, Type 2 diabetes, Vascular disease, Vascular remodeling, Vasculature, Vasculopathy, Ventricular cardiomyocytes, Ventricular fibrillation.

### SUPPLEMENTARY RESULTS

**Clinical and demographic characteristics**

The study population for the meta-analysis consisted of 1,279,610 participants, of which 302,287 were from MVP (43,344 cases and 258,943 controls) and 977,323 were from HERMES Consortium (47,309 cases and 930,014 controls). The clinical and demographic features of the participants are summarized in Table S1. In both the MVP and the HERMES Consortium, the participants with HF were older on average and with a higher prevalence of cardiometabolic risk factors and comorbidities compared to participants without HF (Table S1). The MVP cohort was predominantly male (96.92% of cases and 92.14% of controls, compared to 59.13% of cases and 47.15% of controls in the HERMES Consortium). Atrial fibrillation was present in 34.44% of cases in the MVP and 40.49% of cases in the HERMES Consortium. In the MVP, 69.72% of cases had coronary heart disease, 42.47% of cases had peripheral vascular disease, and 24.93% had stroke. In the HERMES Consortium, 41.90% of cases had myocardial infarction and 60.68% had coronary heart disease. Further, 46.76% cases and 20.61% controls had diabetes in the MVP, compared to 27.05% cases and 6.59% controls in the HERMES Consortium. Detailed breakdown of clinical and demographic characteristics by each study included in the HERMES Consortium has been previously published[5].

**Epigraph Results**

For eight of the hit genes, the wealth of information curated in the EpiGraphDB database[11] provided support of associations with HF-related medical terms. The database identified 77 associations comprising 40 unique medical terms. For every reported association, the EpiGraphDB API provides a list of supporting publications (PubMed ID) and a predicate highlighting the association between the gene and the medical term (e.g., “AFFECTS”, “INHIBITS”, and “AUGMENTS”). These results showed APOC3 affects cardiovascular disease[12] and is associated with diabetes[13–15] and coronary heart disease[16,17]. APOH is associated with dyslipidemias[18] and diabetes[19]. DLL1 is associated with myocardial infarction[20], heart valve diseases[21], and diabetes[22–25], and augments heart diseases[26]. ITIH4 is associated with graft-vs-host diseases[27] and causes endothelial dysfunction[28]. MAPK3 is associated to MI[29] and hypertrophic cardiomyopathy[30], and affects diabetes[31]. SIRPA is associated with diabetes[32] and stimulates vascular endothelial growth factor receptor-2[33]. TNFSF12 is associated with cardiovascular disease[34,35] and is *negatively* associated with heart failure[36]. TNXB interacts with vascular endothelial growth factor B[37] and is associated with chronic kidney failure[38]. The complete list of the predicates is provided in the Supplementary Data.

### Supplementary Figures

Supplementary Figure 1. Quantile-quantile (Q-Q) plot of HF GWAS.

Supplementary Figure 2. Locus zoom plots for the eighteen novel HF GWAS-associated loci.

Supplementary Figure 3. Genetic associations on heart failure and heart failure risk factors and cardiac MRI traits.

Supplementary Figure 4. Genetic associations of 18 HF loci against cardiac MRI traits.

Supplementary Figure 5. Pipeline of analyses for assessment of horizontal pleiotropy.

Supplementary Figure 6. APOH PPI Network.

Supplementary Figure 7. Pathway enrichment analysis of HF variants identified through GWAS and MR proteomics.

Supplementary Figure 8. Downstream enrichment analysis of 18 new HF GWAS variants.

**Supplementary Figure 1. Quantile-quantile (Q-Q) plot of HF GWAS**. The Q-Q plot demonstrates the number and magnitude of observed associations between genotyped SNPs and heart failure, compared to the expected association statistics under the null hypothesis that there is no association. The identity line is shown in red. Observed association statistics (y-axis) and expected association statistics (x-axis) are on a $-log_{10}P$ scale.

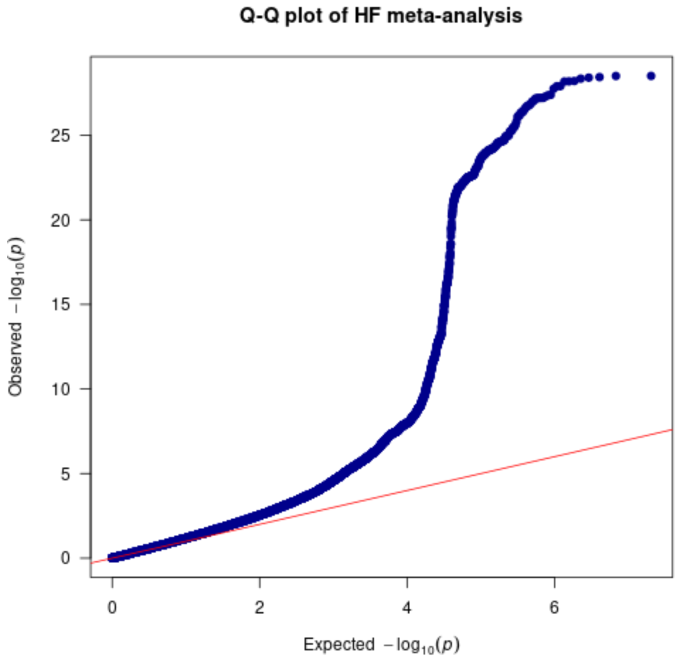

**Supplementary Figure 2. Locus zoom plots for the eighteen novel HF GWAS-associated loci**. The plots illustrate fine mapping of the regions flanking each of the eighteen novel HF GWAS-associated loci. The color coding indicates linkage disequilibrium of each SNP as measured by $r^2$ according to the shown scale with the relevant GWAS SNP annotated.

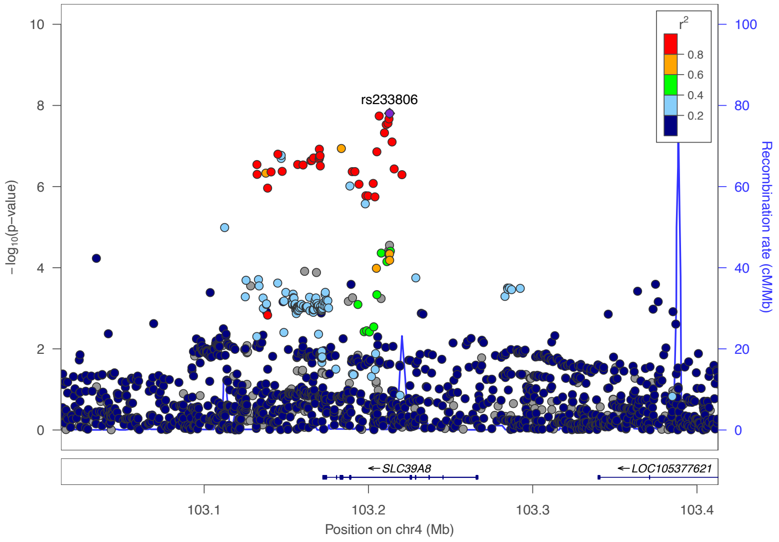


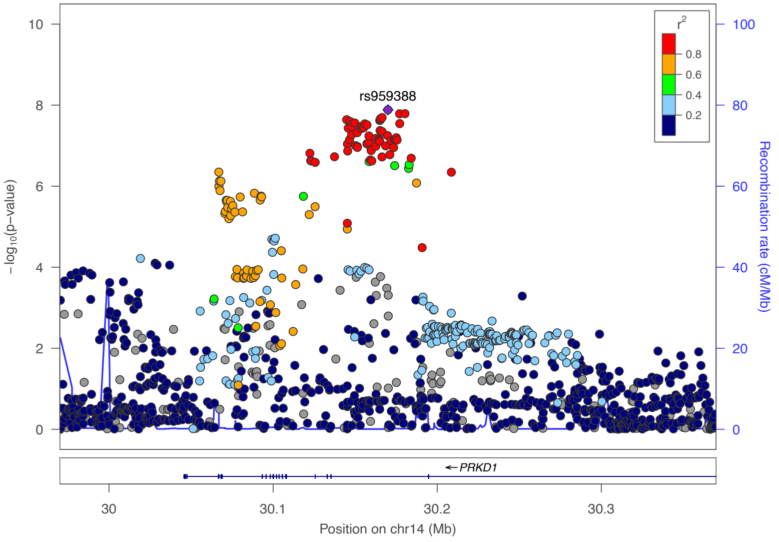


**Supplementary Figure 2a.** Locus zoom plot for rs233806.

**Supplementary Figure 2b**. Locus zoom plot for rs959388.

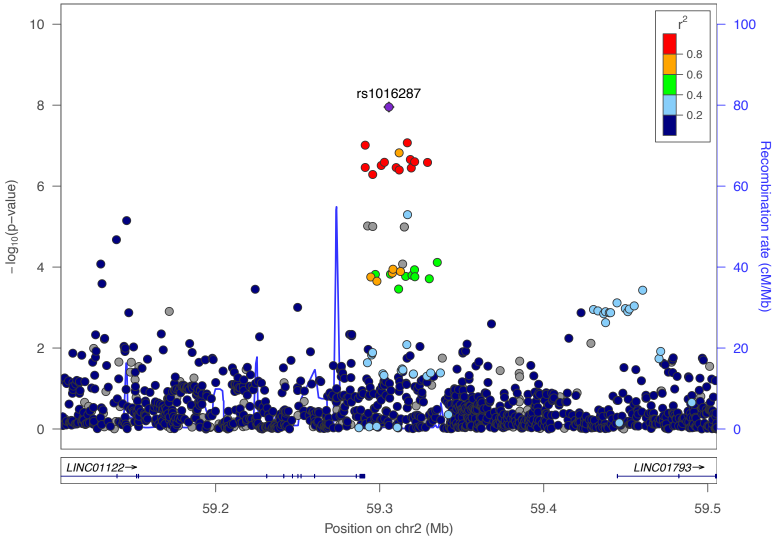


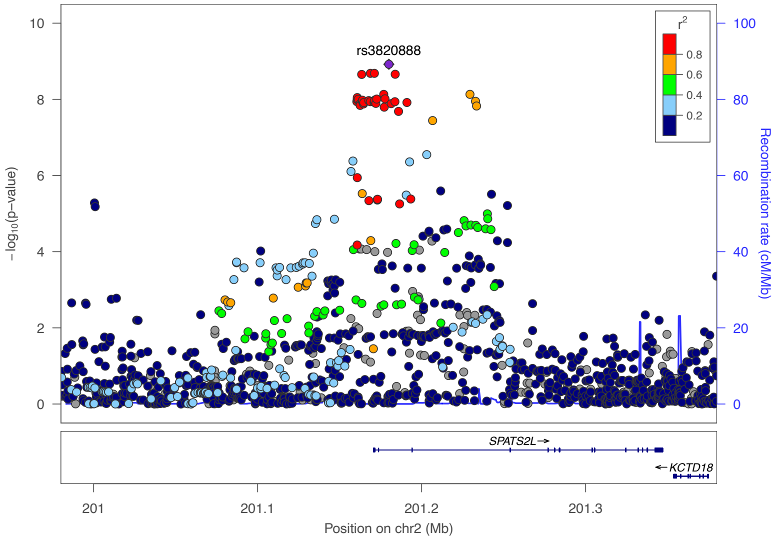


**Supplementary Figure 2c**. Locus zoom plot for rs1016287.

**Supplementary Figure 2d.** Locus zoom plot for rs3820888

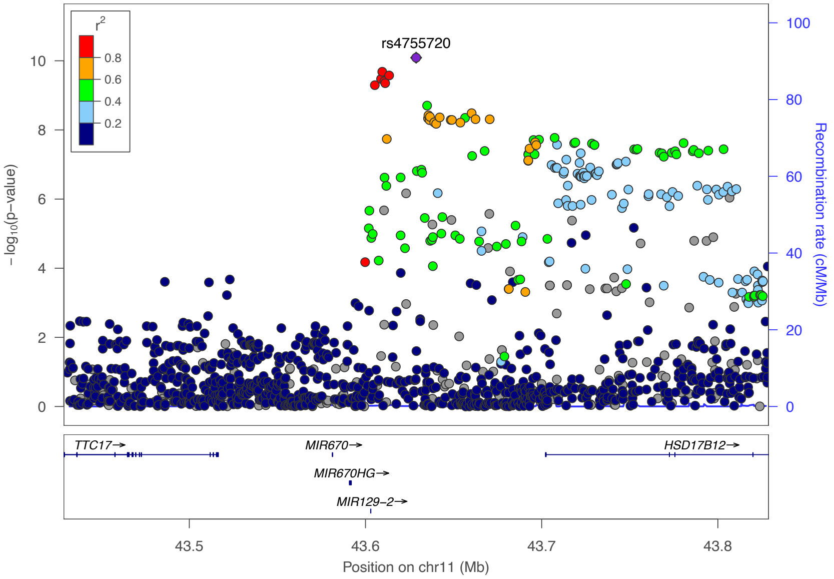


**Supplementary Figure 2e.** Locus zoom plot for rs4755720.

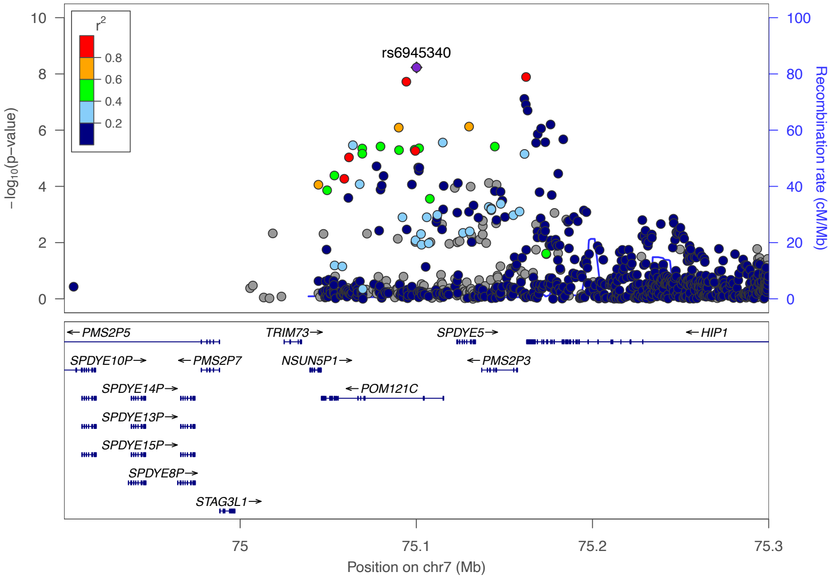


**Supplementary Figure 2f.** Locus zoom plot for rs6945340.

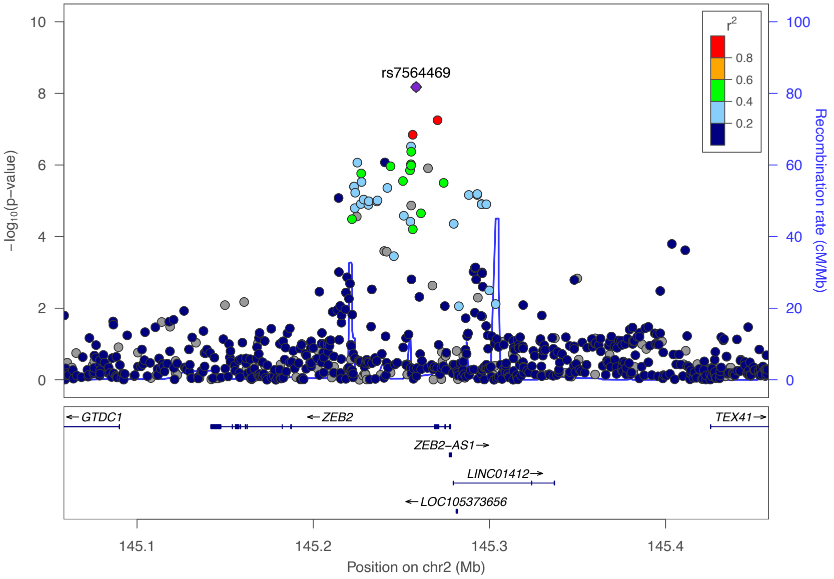


**Supplementary Figure 2g.** Locus zoom plot for rs7564469.

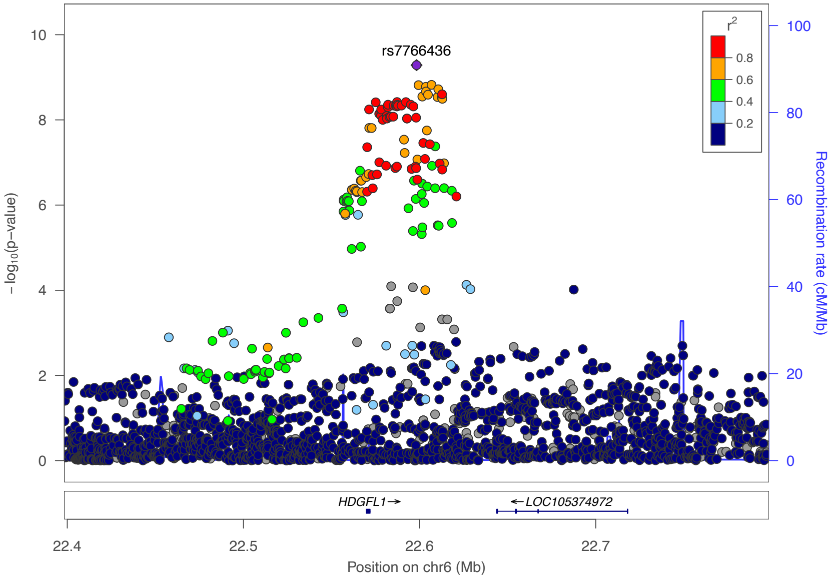


**Supplementary Figure 2h.** Locus zoom plot for rs7766436.

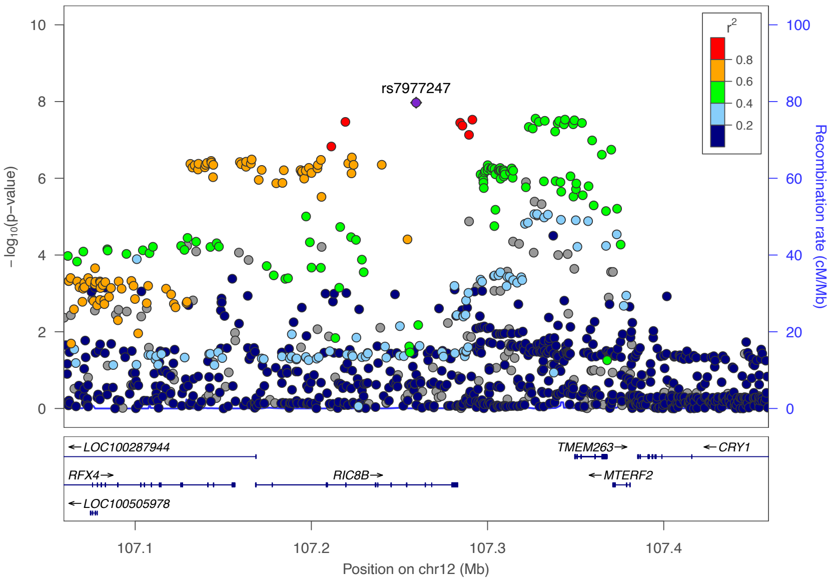


**Supplementary Figure 2i**. Locus zoom plot for rs7977247.

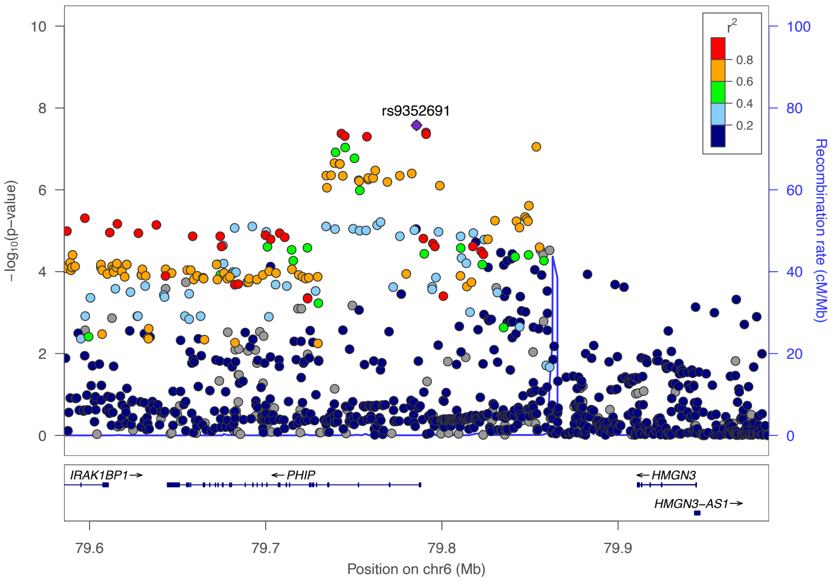


**Supplementary Figure 2j**. Locus zoom plot for rs9352691.

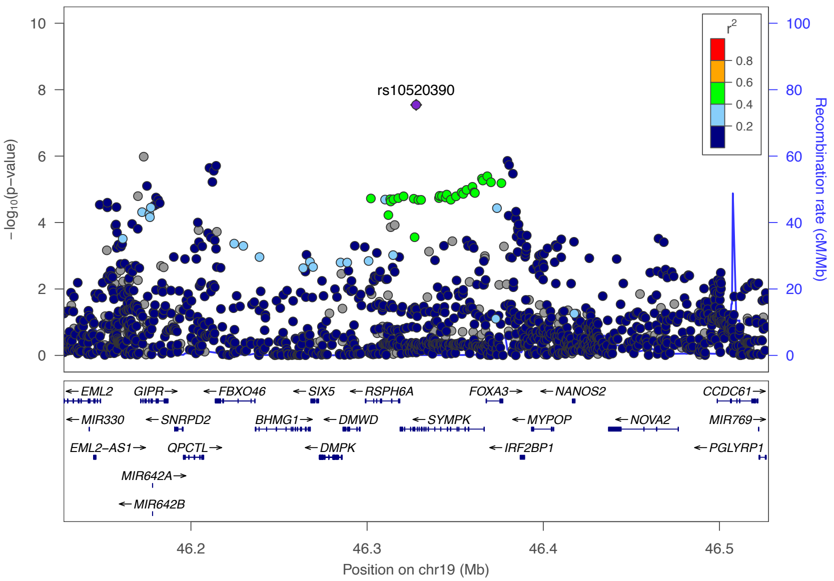


**Supplementary Figure 2k.** Locus zoom plot for rs10520390.

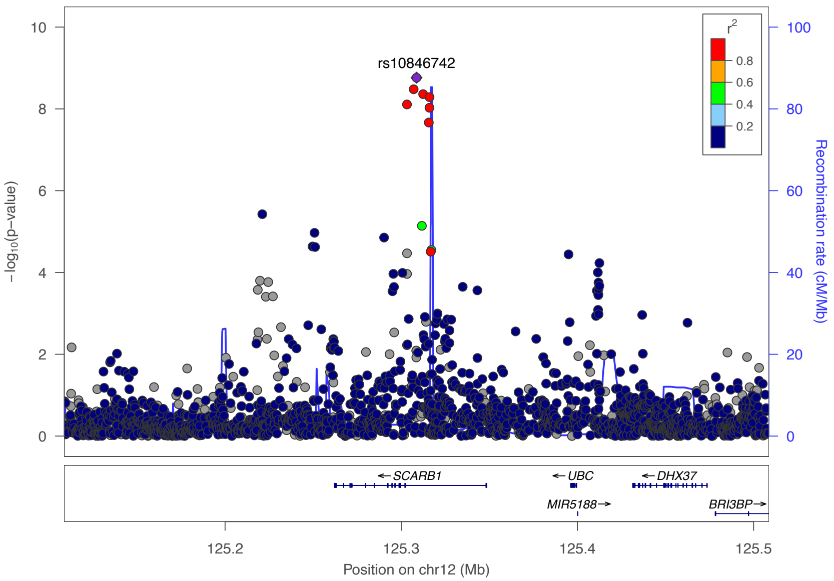


**Supplementary Figure 2l**. Locus zoom plot for rs10846742.

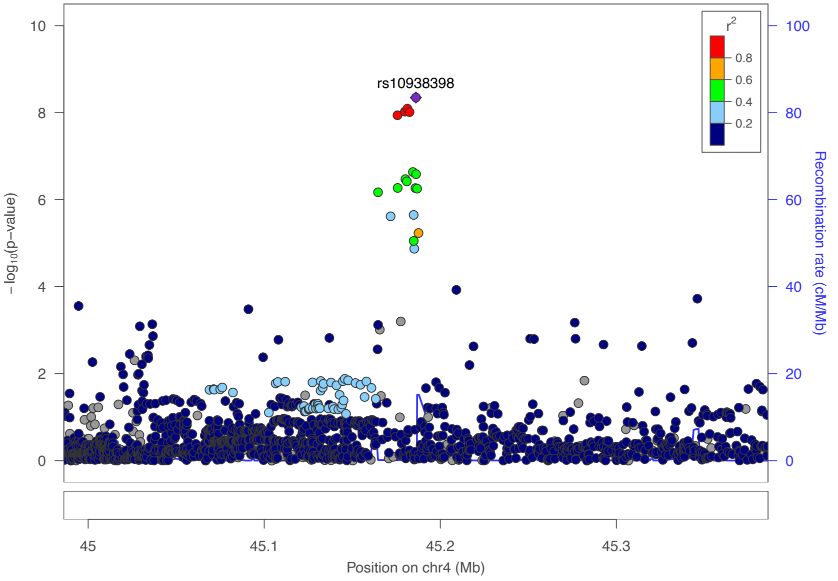


**Supplementary Figure 2m.** Locus zoom plot for rs10938398.

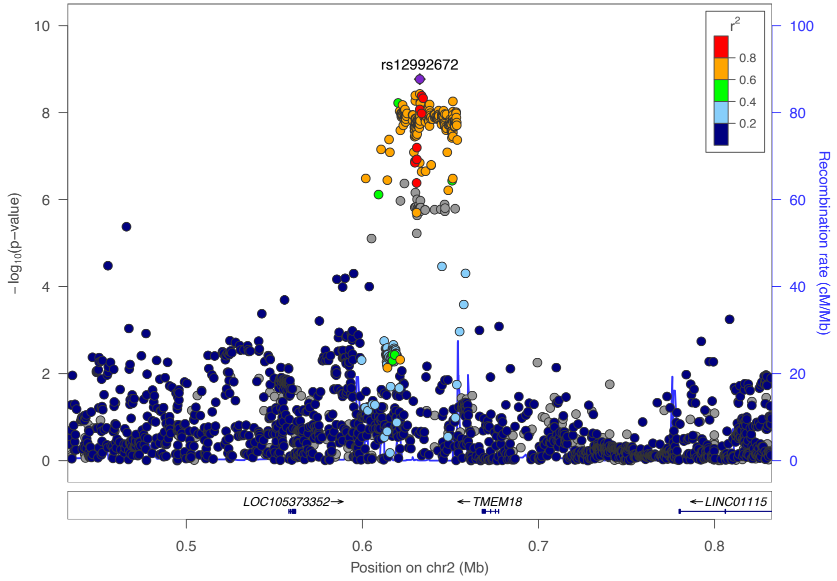


**Supplementary Figure 2n.** Locus zoom plot for rs12992672.

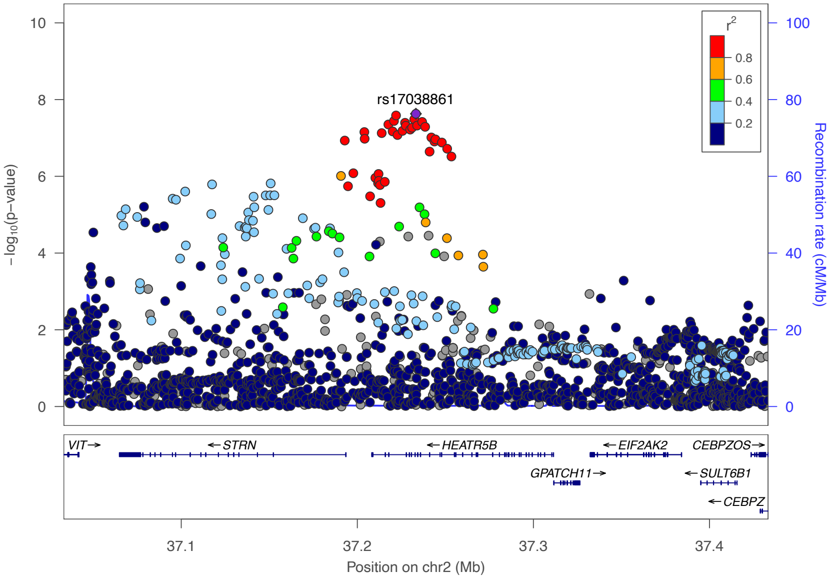


**Supplementary Figure 2o.** Locus zoom plot for rs17038861.

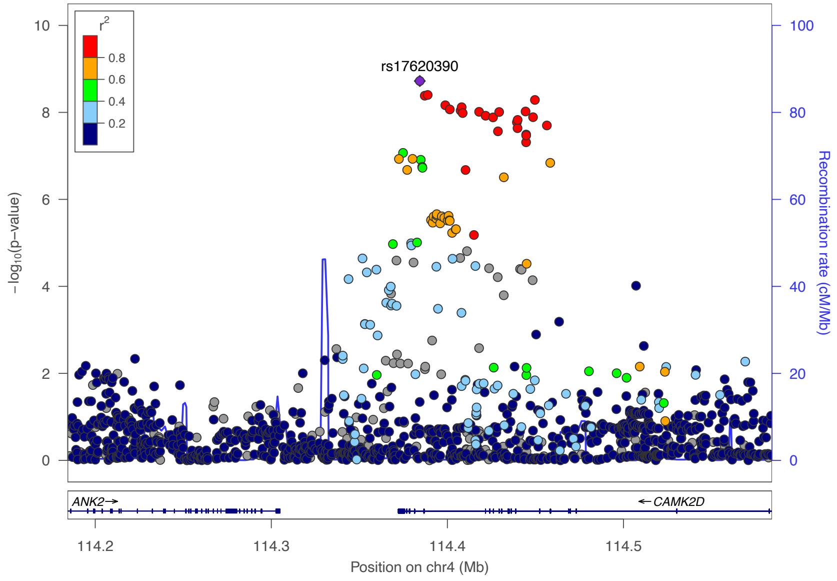


**Supplementary Figure 2p.** Locus zoom plot for rs17620390.

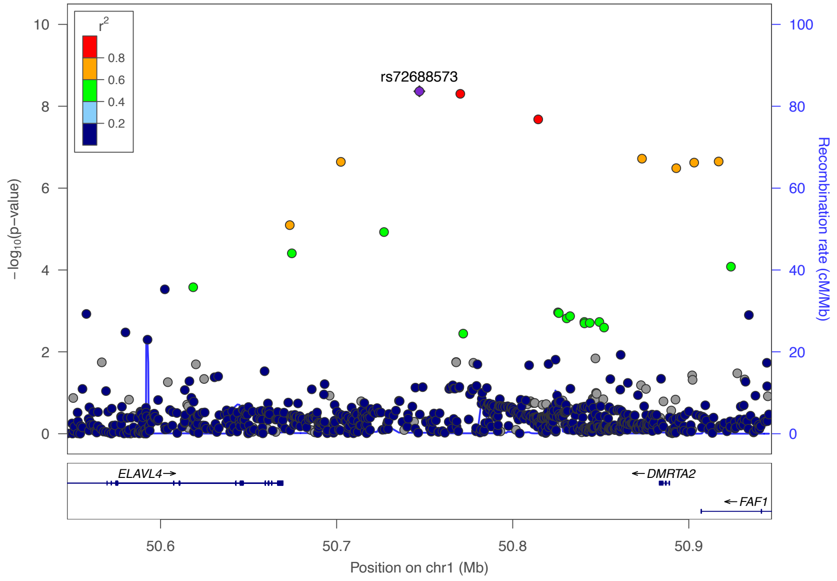


**Supplementary Figure 2q.** Locus zoom plot for rs72688573.

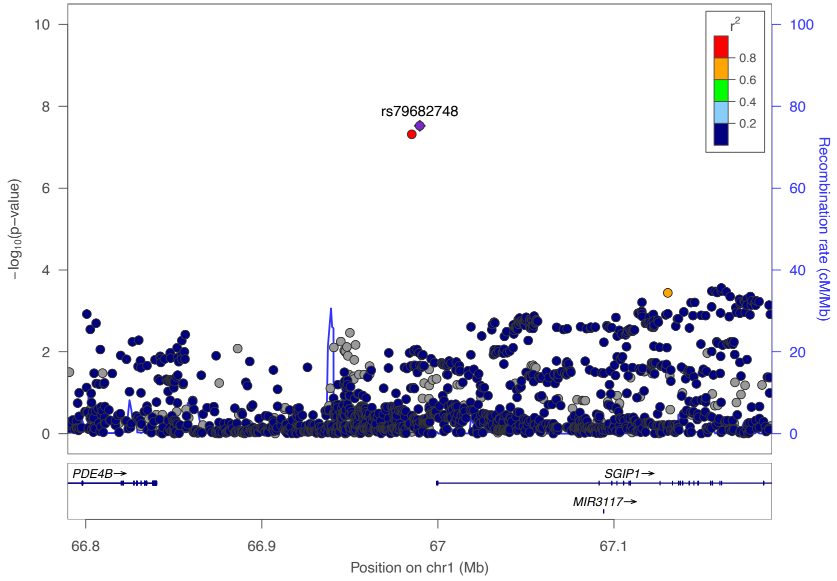


**Supplementary Figure 2r**. Locus zoom plot for rs79682748.

**Supplementary Figure 3. Genetic associations on heart failure and heart failure risk factors and cardiac MRI traits.** n = 1,279,610 participants for heart failure, n = 681,275 for body mass index, n = 414,343 for alcohol consumption, n = 1,030,836 for atrial fibrillation, n = 757,601 for diastolic blood pressure, n = 757,601 for systolic blood pressure, n = 655,666 for type 2 diabetes, n = 547,261 for coronary artery disease, n = 1,320,016 for low-density lipoprotein cholesterol, n = 1,320,016 for high-density lipoprotein cholesterol, n = 1,201,909 for estimated glomerular filtration rate (eGFR), n = 462,933 for chronic obstructive airways disease, n = 3,301 for troponin I cardiac muscle, n = 3,301 for NT-PBNP, n = 21,758 for IL-6, n = 36,041 for cardiac MRI traits (left ventricular end-diastolic volume, left ventricular end-diastolic volume BSA-indexed, left ventricular ejection fraction, left ventricular end-systolic volume, left ventricular end-systolic volume-BSA indexed, stroke volume, stroke volume BSA-indexed, left ventricular mass, and left ventricular mass to end-diastolic volume ratio). The measure of centre for the error bars is the effect estimate (beta coefficient) of the genetic association between the loci (specified in each figure legend) and trait. The error bars represent the 95% confidence interval around the effect estimate. All tests were two-sided without adjustment for multiple comparisons.

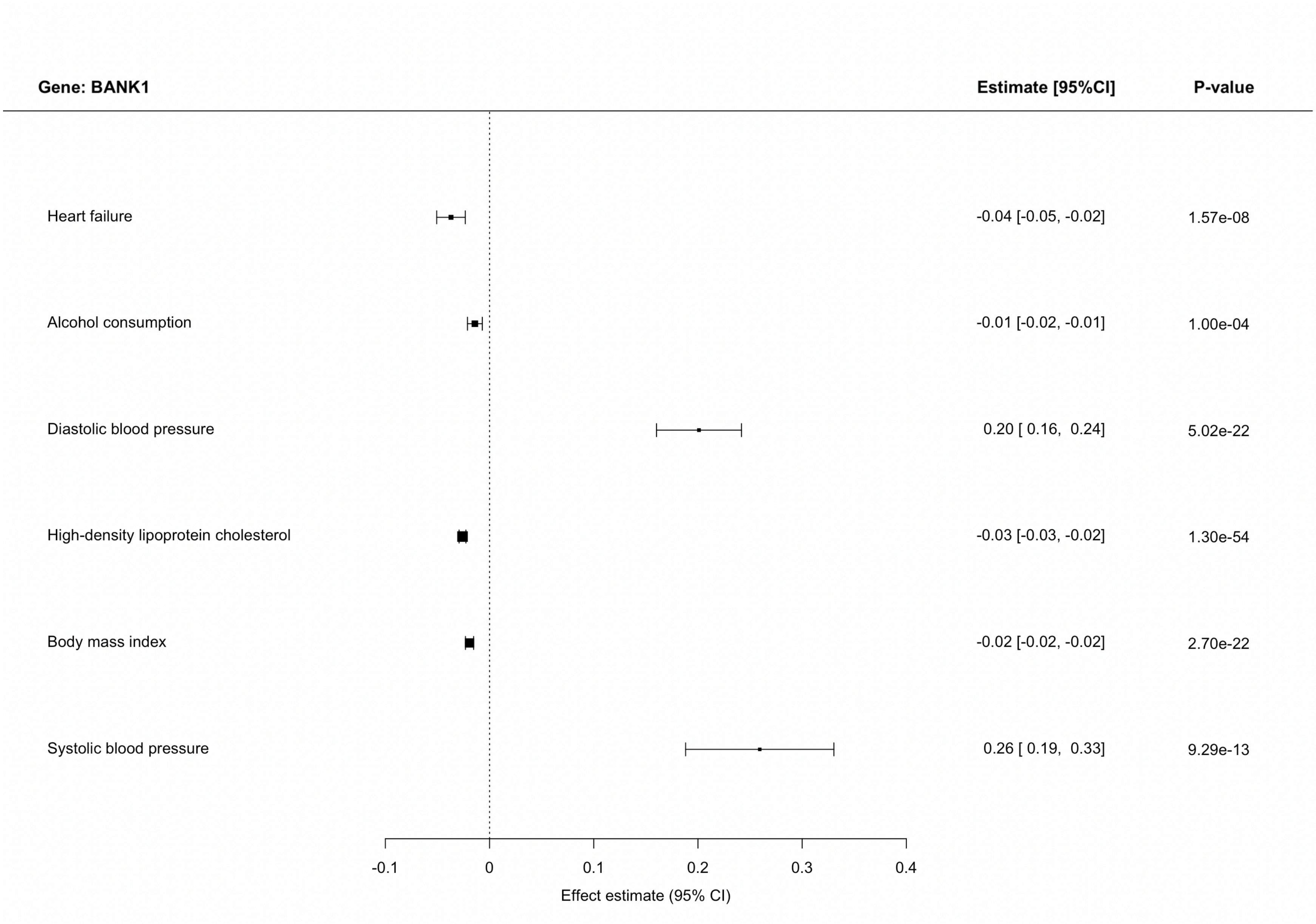


**Supplementary Figure 3a.** Genetic associations on heart failure and heart failure risk factors and cardiac MRI traits that pass a threshold of p<0.0001 for rs233806 (BANK1).

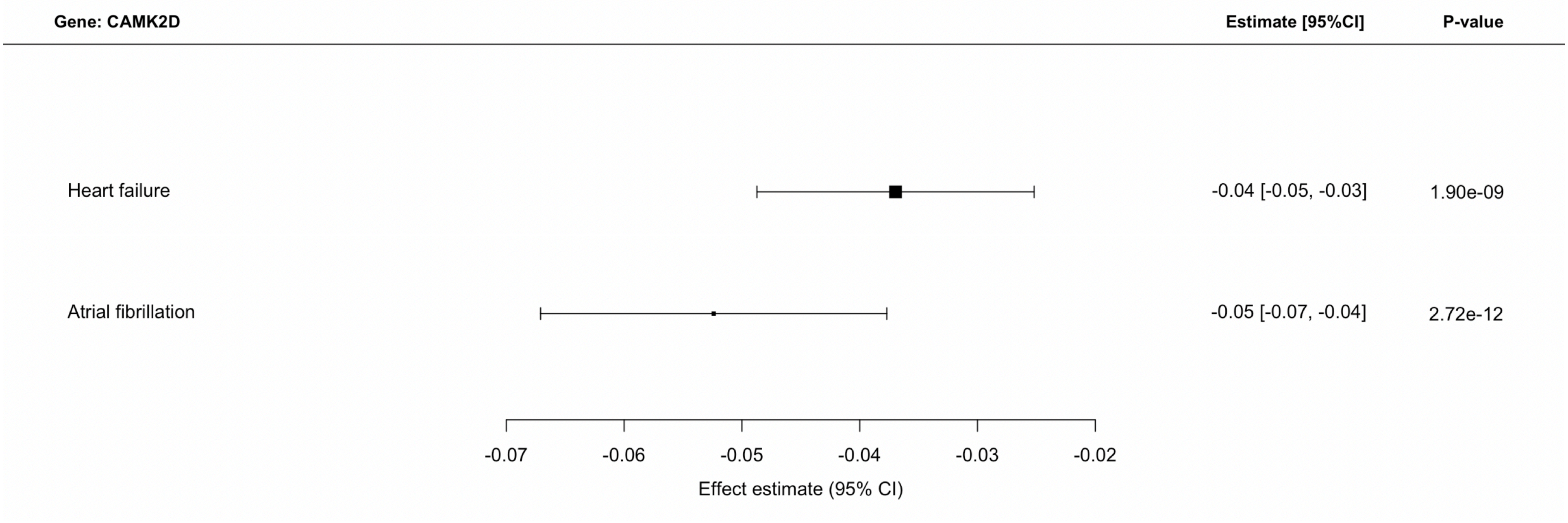


**Supplementary Figure 3b.** Genetic associations on heart failure and heart failure risk factors and cardiac MRI traits that pass a threshold of p<0.0001 for rs17620390 (CAMK2D).

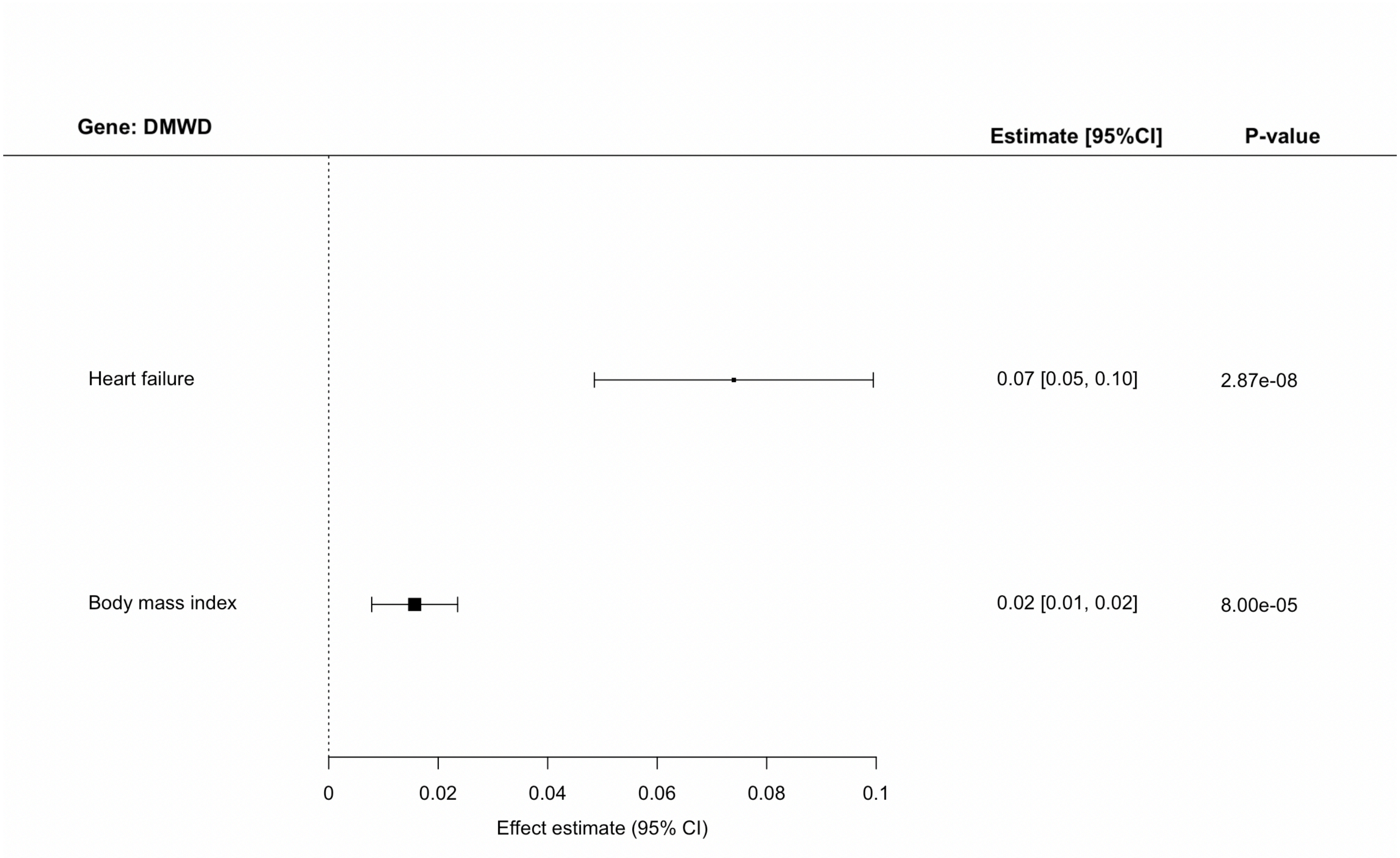


**Supplementary Figure 3c.** Genetic associations on heart failure and heart failure risk factors and cardiac MRI traits that pass a threshold of p<0.0001 for rs10520390 (DMWD).

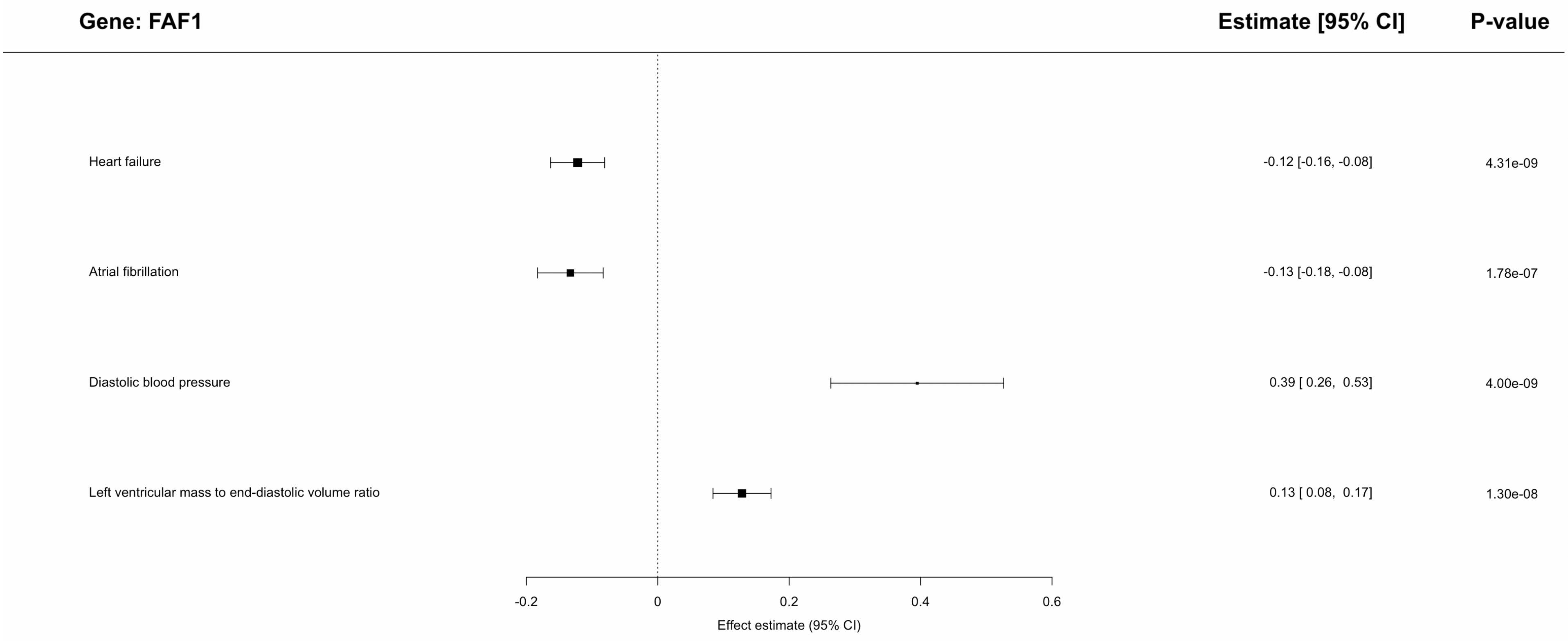


**Supplementary Figure 3d.** Genetic associations on heart failure and heart failure risk factors and cardiac MRI traits that pass a threshold of p<0.0001 for rs72688573 (FAF1).

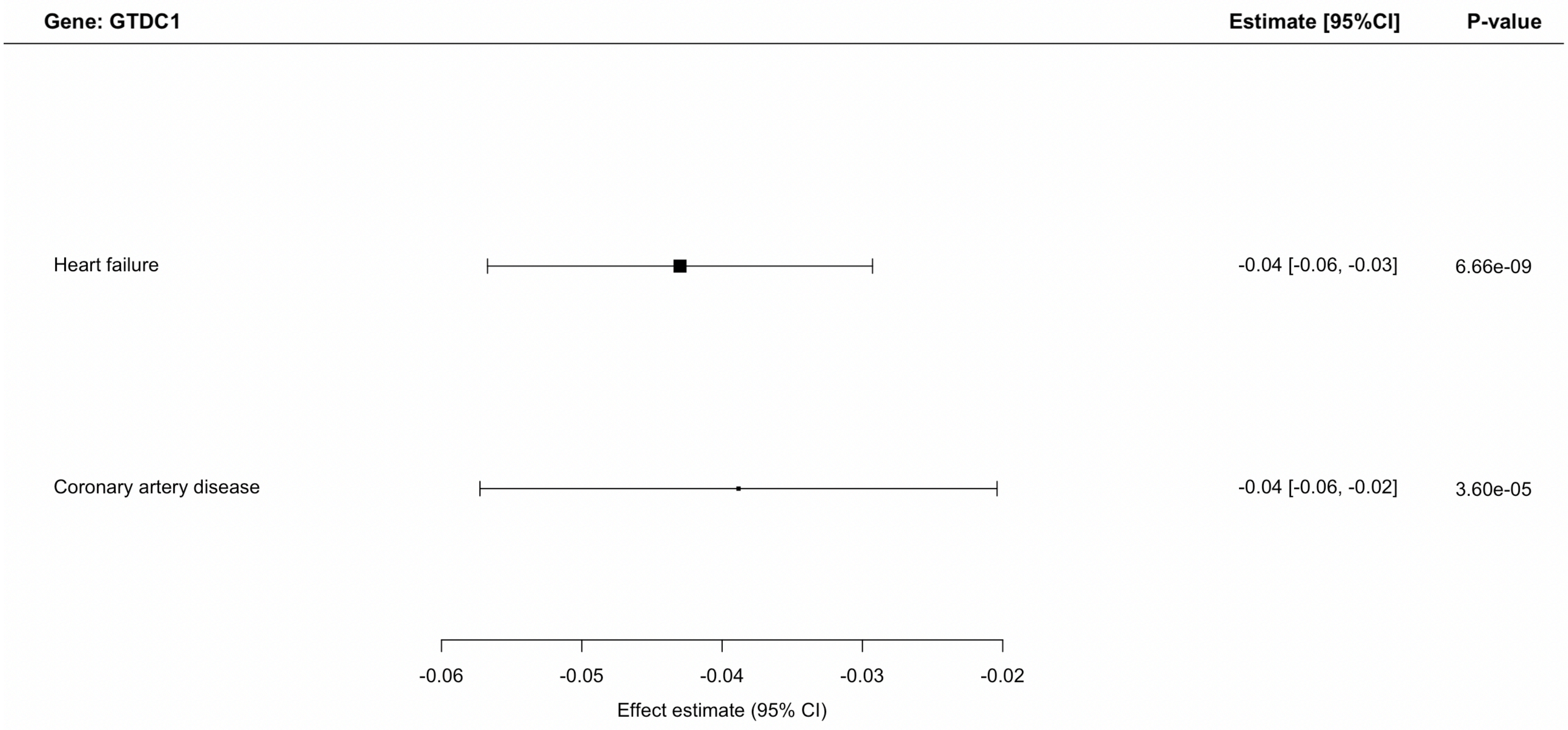


**Supplementary Figure 3e.** Genetic associations on heart failure and heart failure risk factors and cardiac MRI traits that pass a threshold of p<0.0001 for rs7564469 (GTDC1).

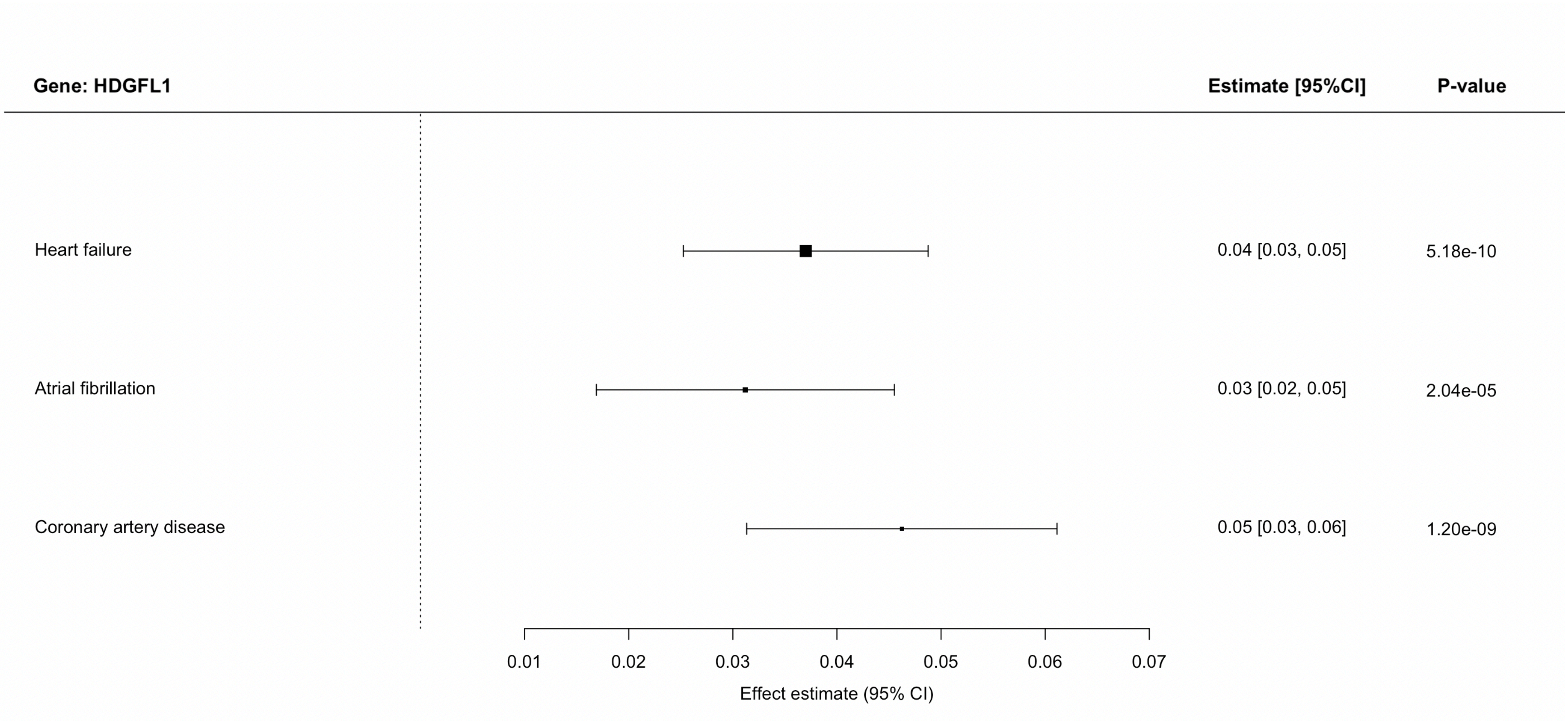


**Supplementary Figure 3f.** Genetic associations on heart failure and heart failure risk factors and cardiac MRI traits that pass a threshold of p<0.0001 for rs7766436 (HDGFL1).

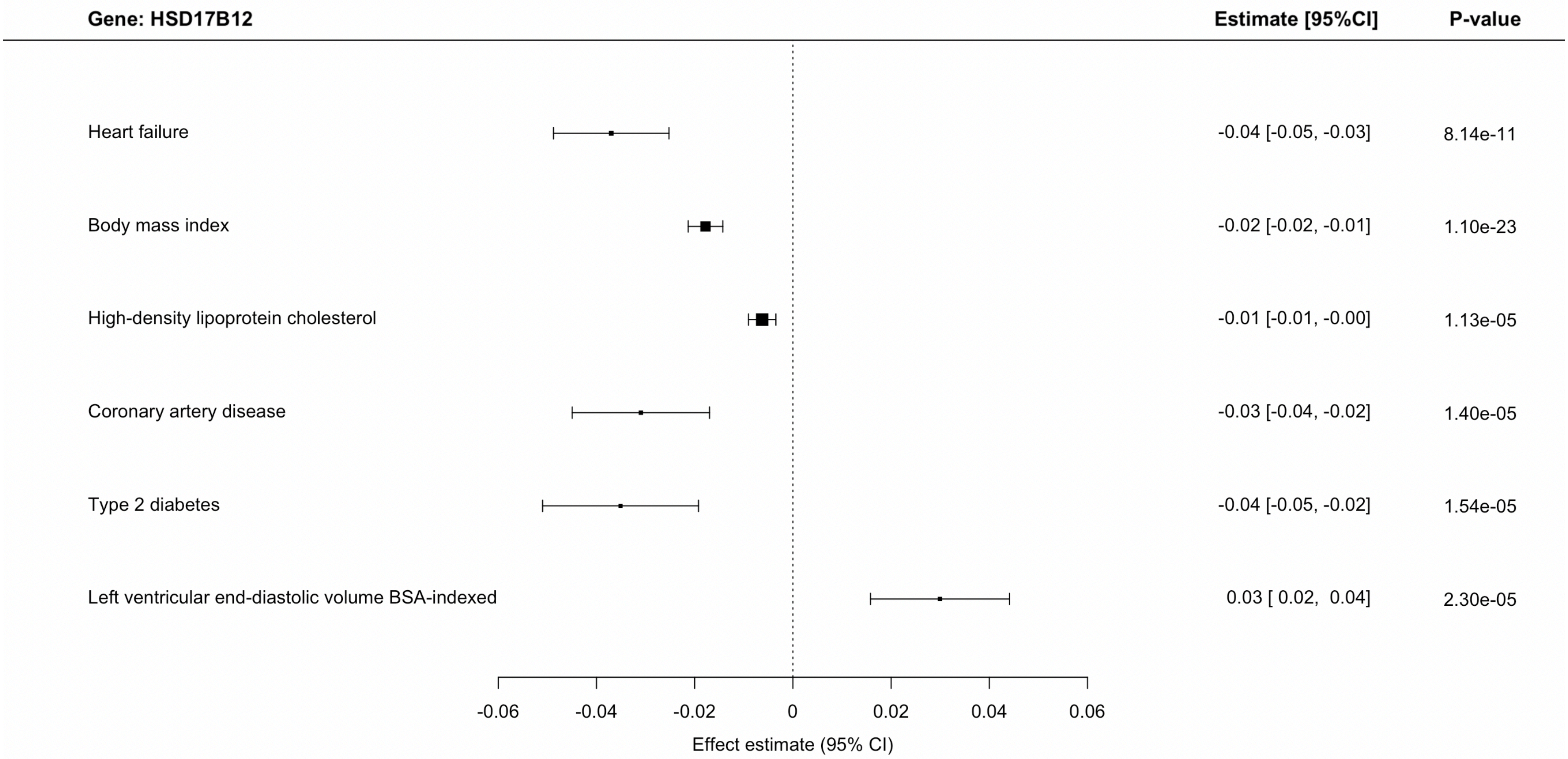


**Supplementary Figure 3g.** Genetic associations on heart failure and heart failure risk factors and cardiac MRI traits that pass a threshold of p<0.0001 for rs4755720 (HSD17B12).

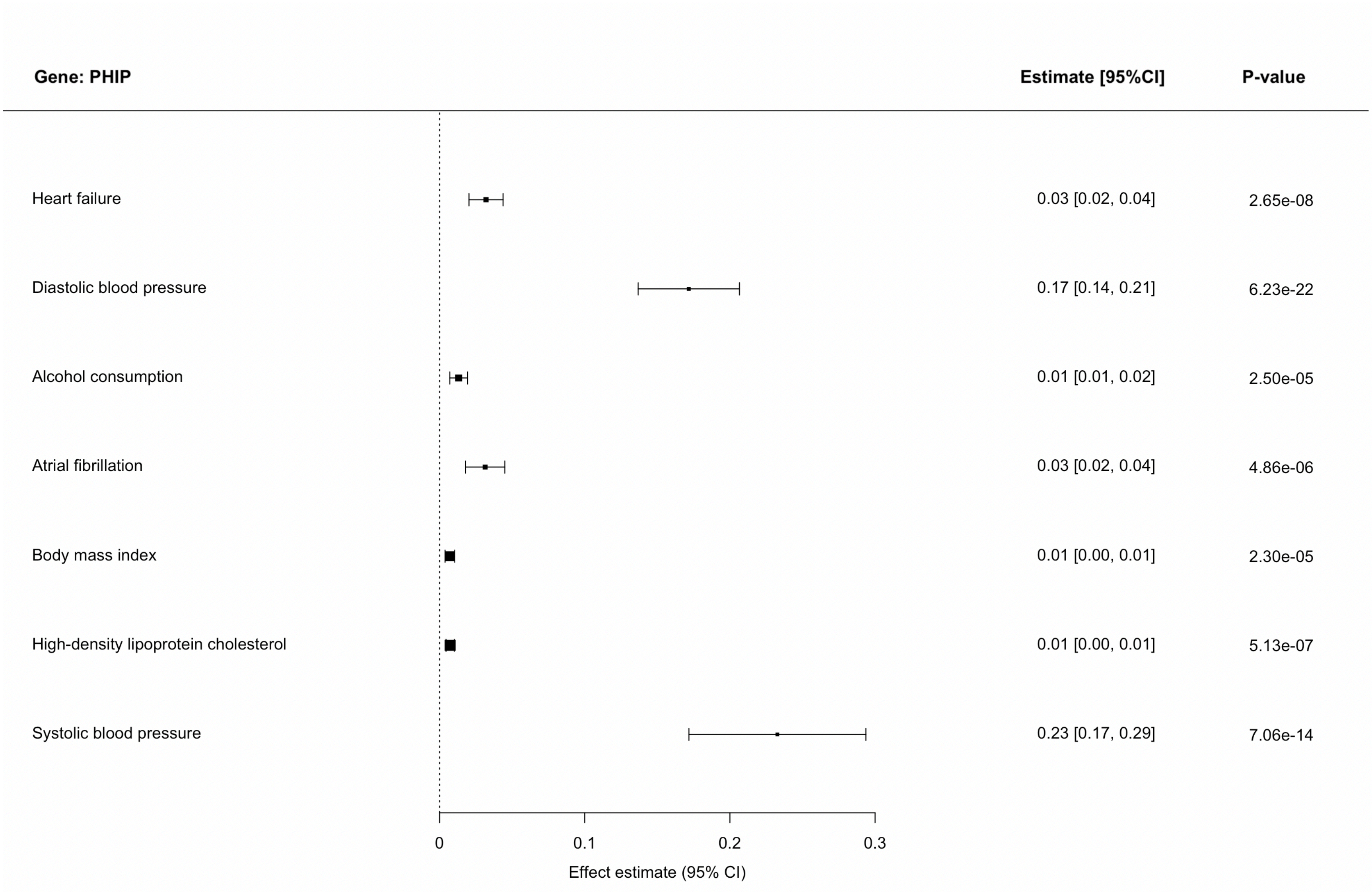


**Supplementary Figure 3h.** Genetic associations on heart failure and heart failure risk factors and cardiac MRI traits that pass a threshold of $p<0.0001$ for rs9352691 (PHIP).

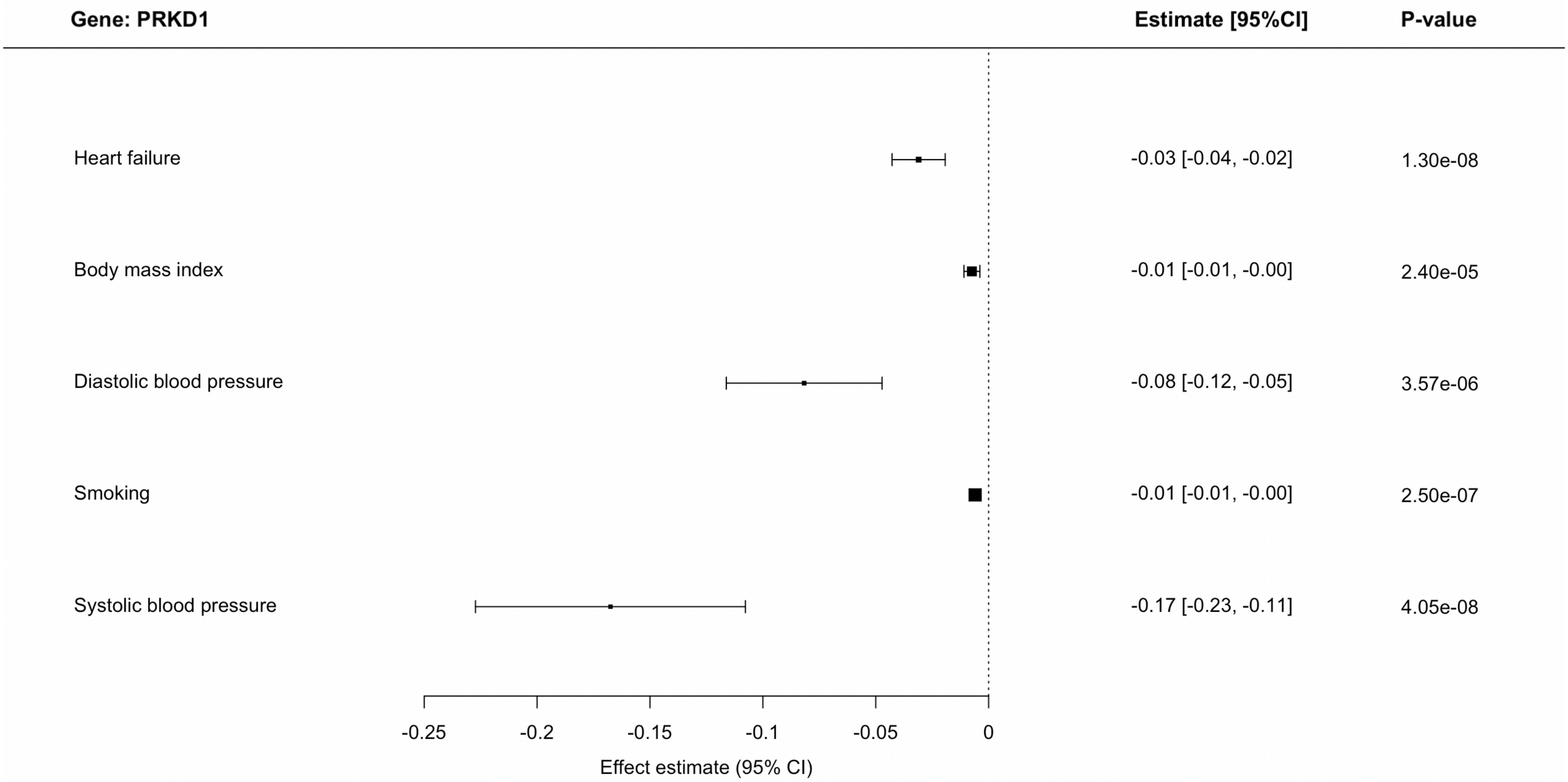


**Supplementary Figure 3i.** Genetic associations on heart failure and heart failure risk factors and cardiac MRI traits that pass a threshold of p<0.0001 for rs959388(PRKD1).

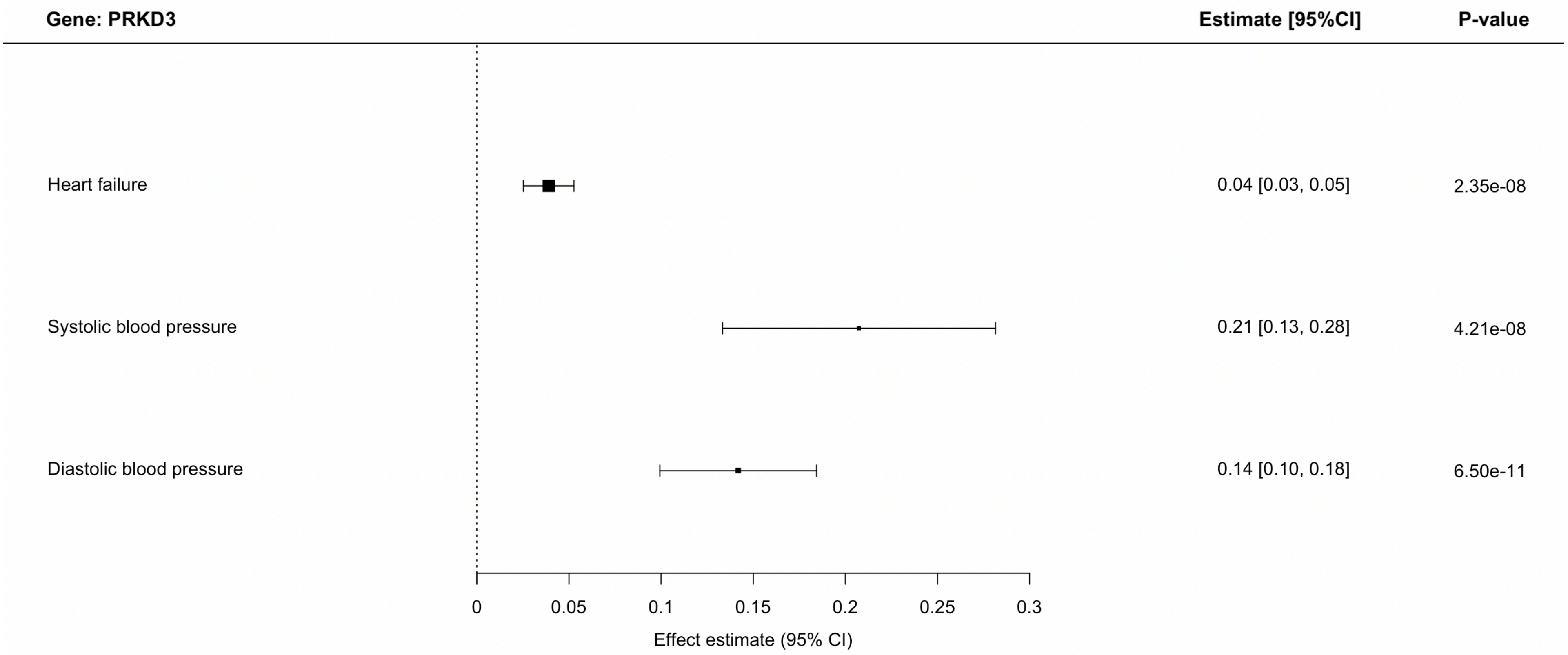


**Supplementary Figure 3j.** Genetic associations on heart failure and heart failure risk factors and cardiac MRI traits that pass a threshold of p<0.0001 for rs17038861 (PRKD3).

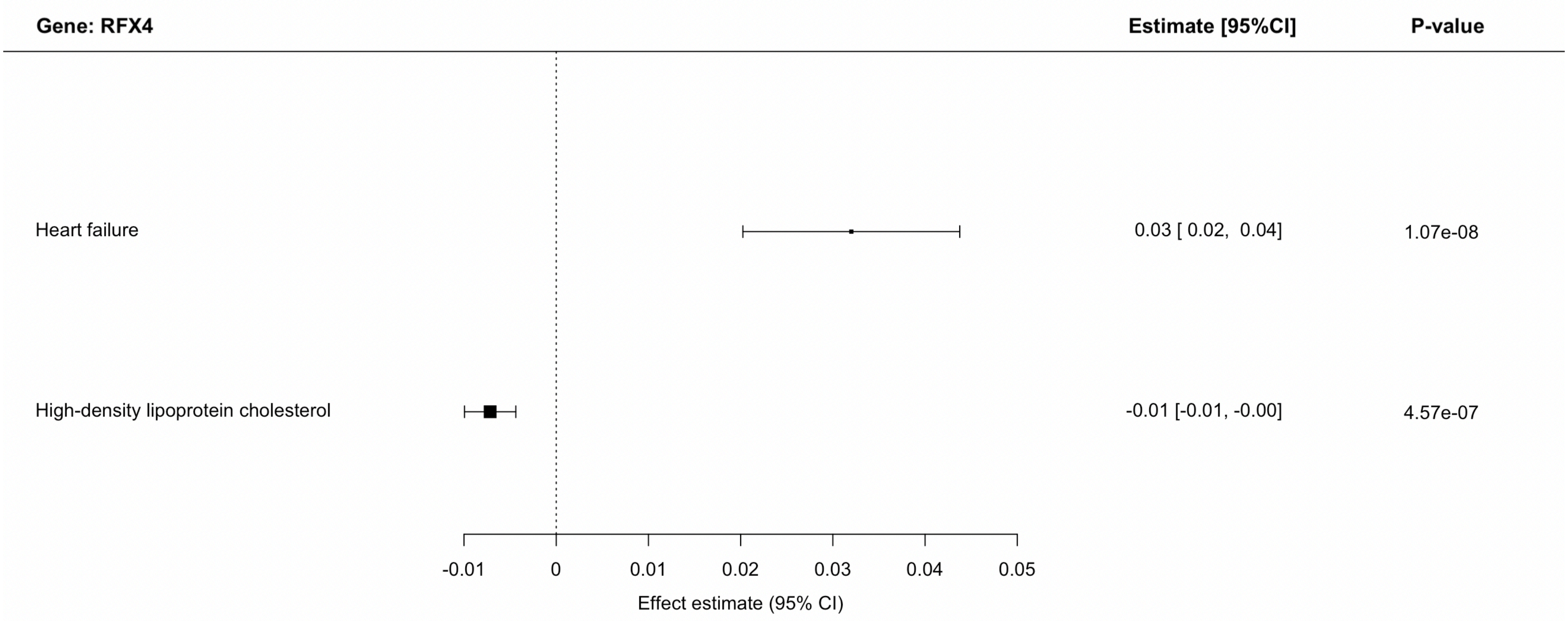


**Supplementary Figure 3k.** Genetic associations on heart failure and heart failure risk factors and cardiac MRI traits that pass a threshold of p<0.0001 for rs7977247 (RFX4).

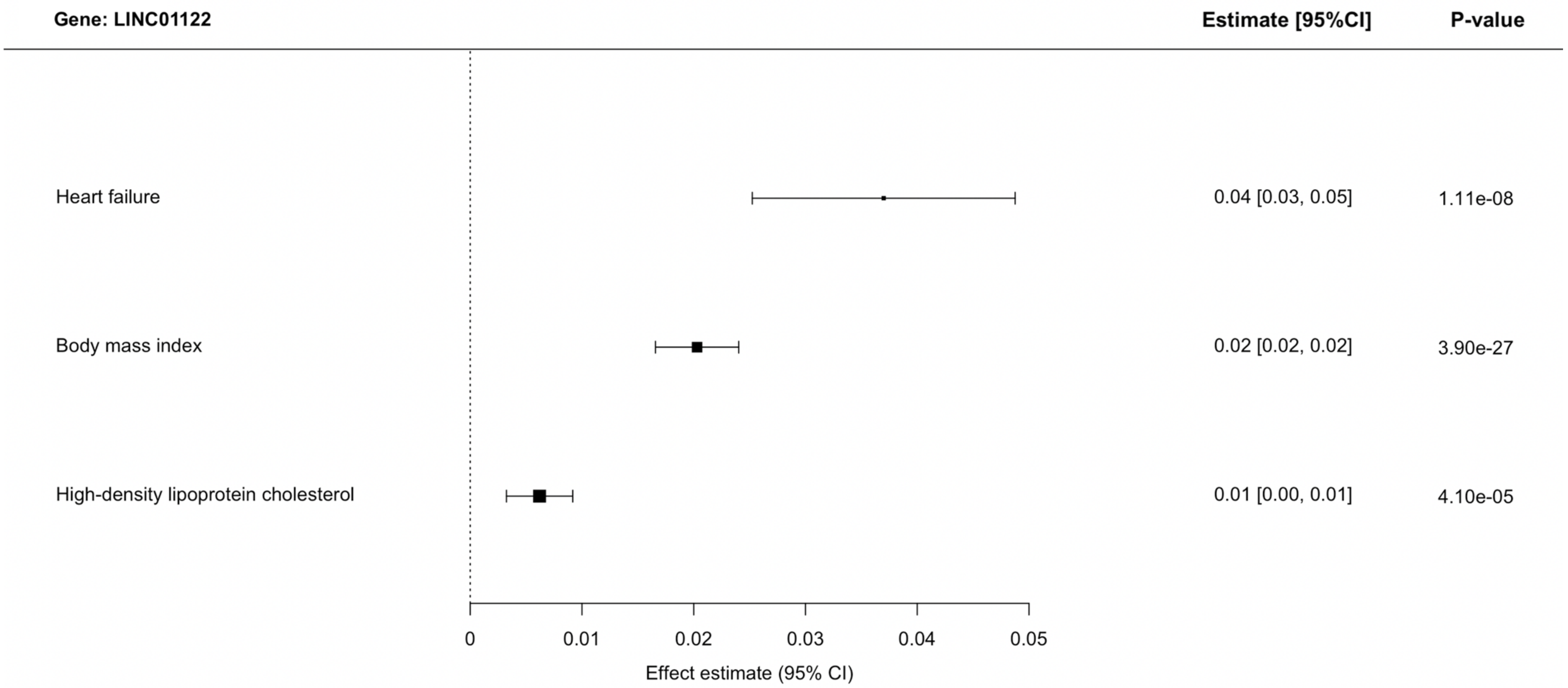


**Supplementary Figure 3l.** Genetic associations on heart failure and heart failure risk factors and cardiac MRI traits that pass a threshold of p<0.0001 for rs1016287 (LINC01122).

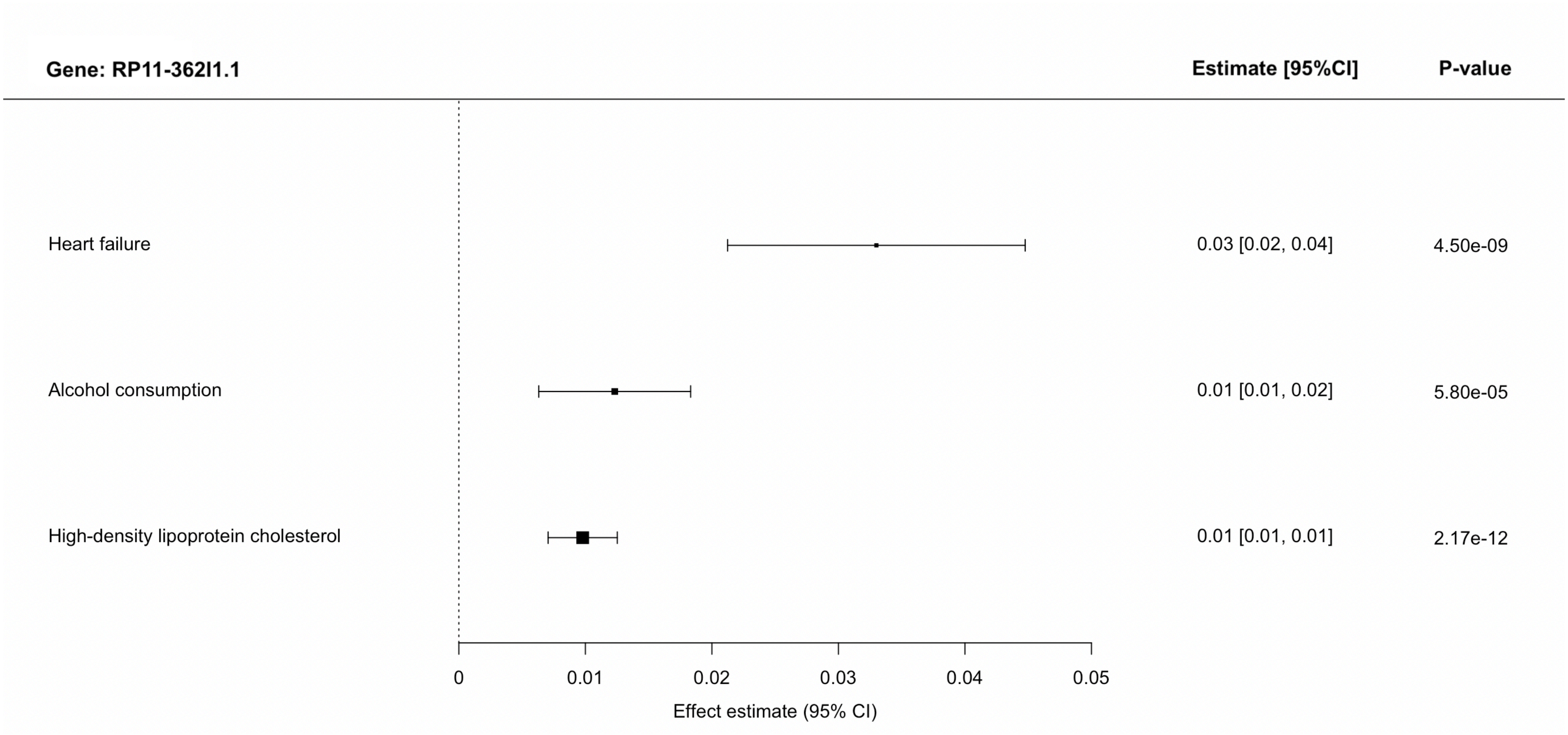


**Supplementary Figure 3m.** Genetic associations on heart failure and heart failure risk factors and cardiac MRI traits that pass a threshold of p<0.0001 for rs10938398 (RP11-362I1.1).

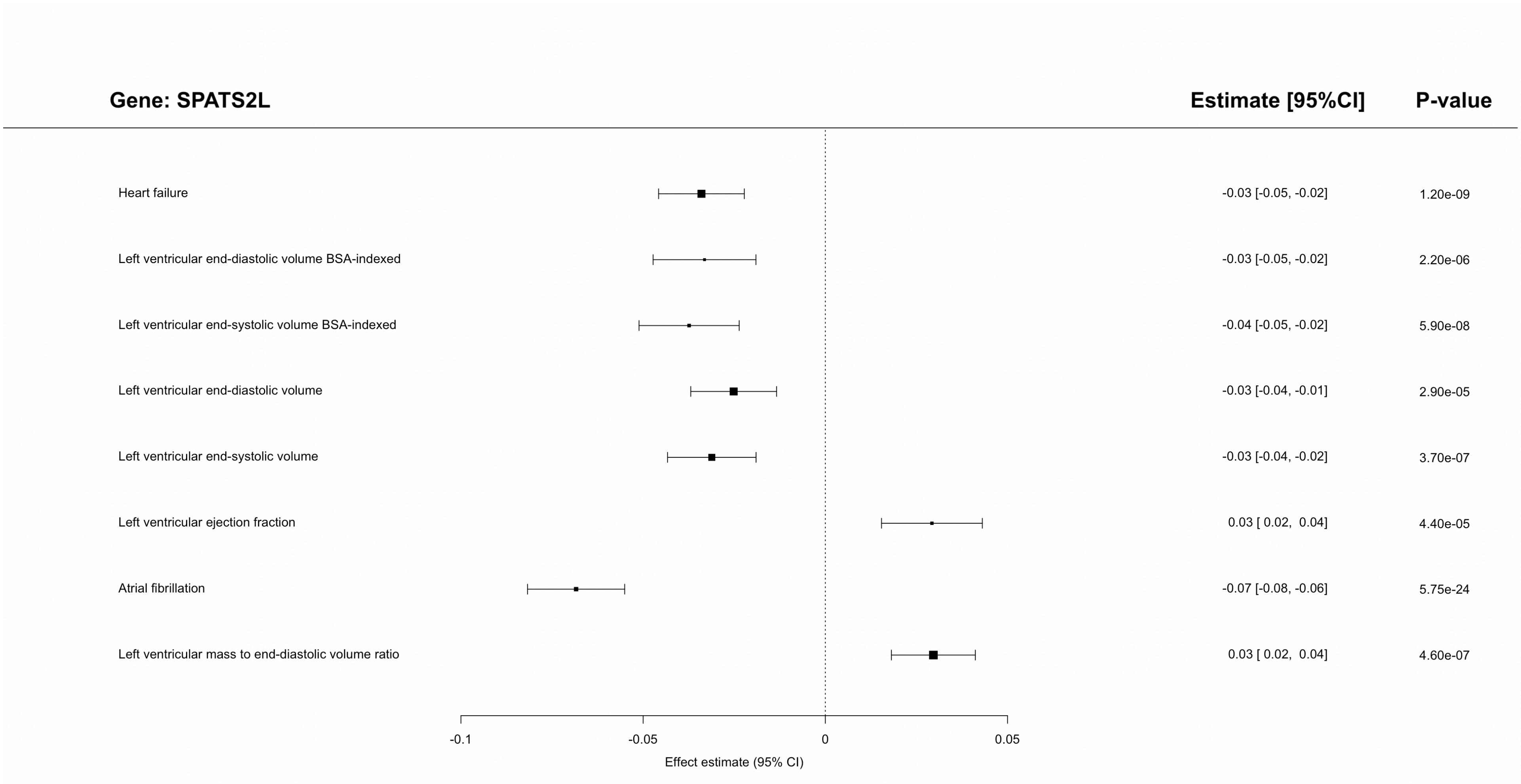


**Supplementary Figure 3n.** Genetic associations on heart failure and heart failure risk factors and cardiac MRI traits that pass a threshold of p<0.0001 for rs3820888 (SPATS2L).

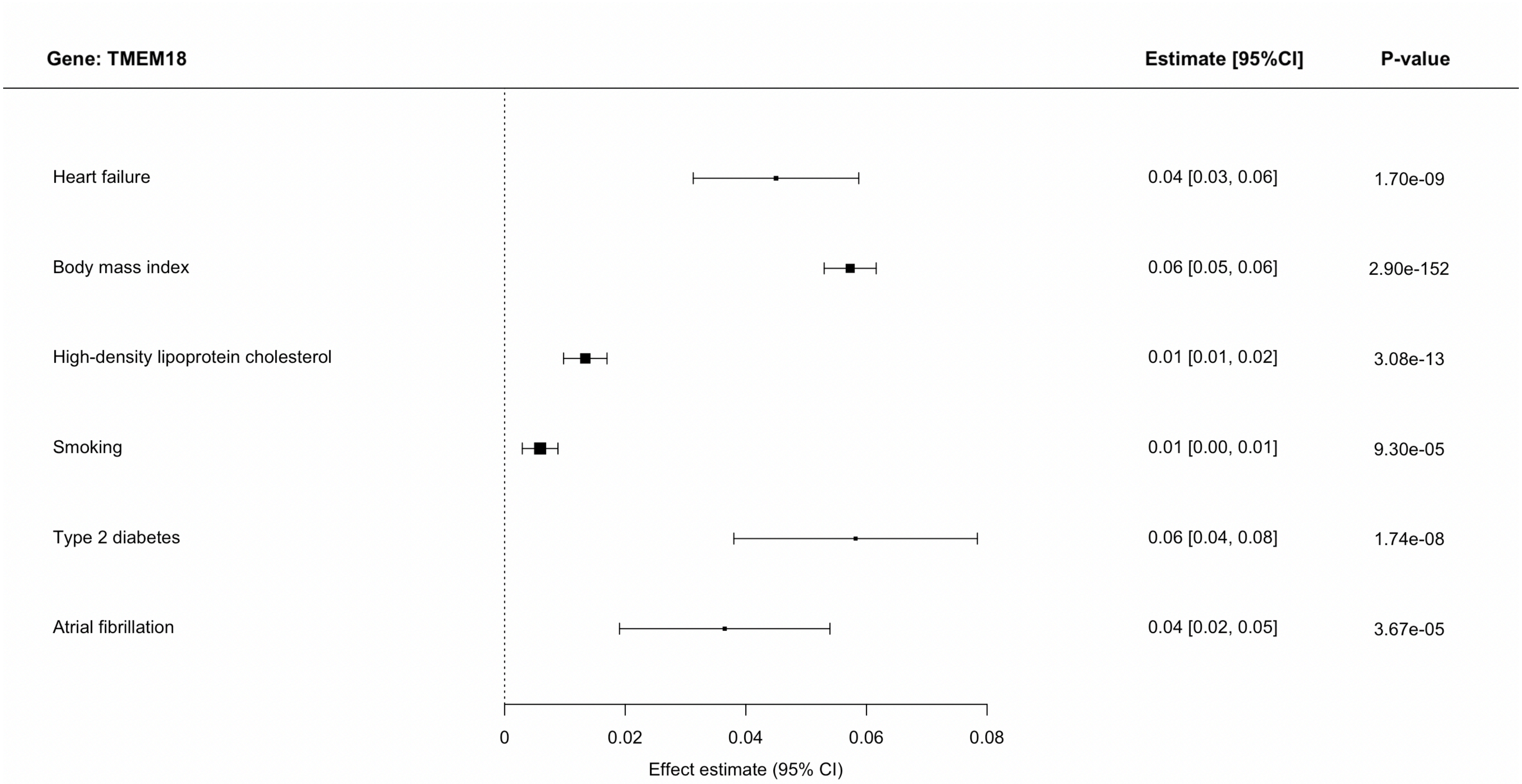


**Supplementary Figure 3o.** Genetic associations on heart failure and heart failure risk factors and cardiac MRI traits that pass a threshold of p<0.0001 for rs12992672 (TMEM18).

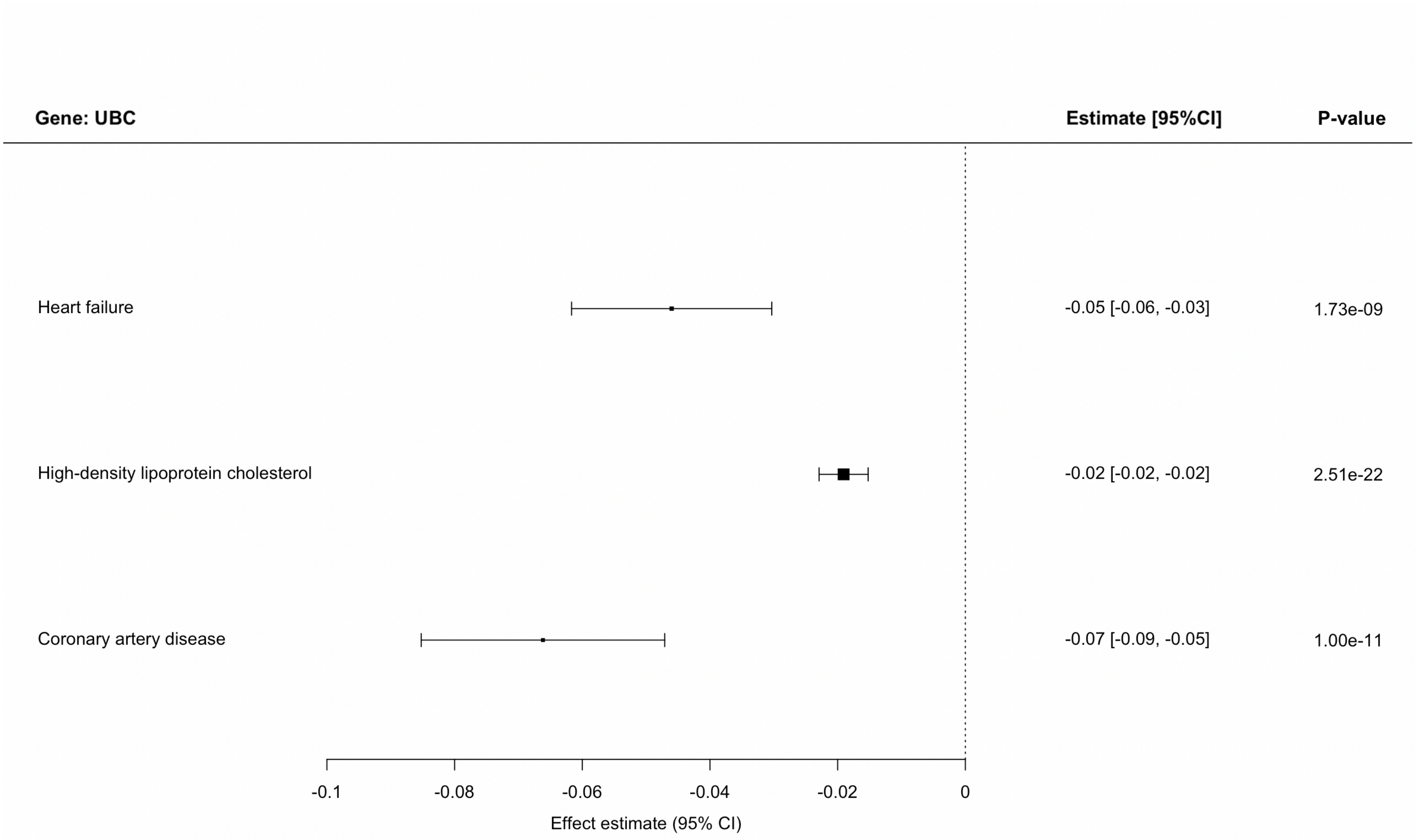


**Supplementary Figure 3p.** Genetic associations on heart failure and heart failure risk factors and cardiac MRI traits that pass a threshold of p<0.0001 for rs10846742 (UBC).

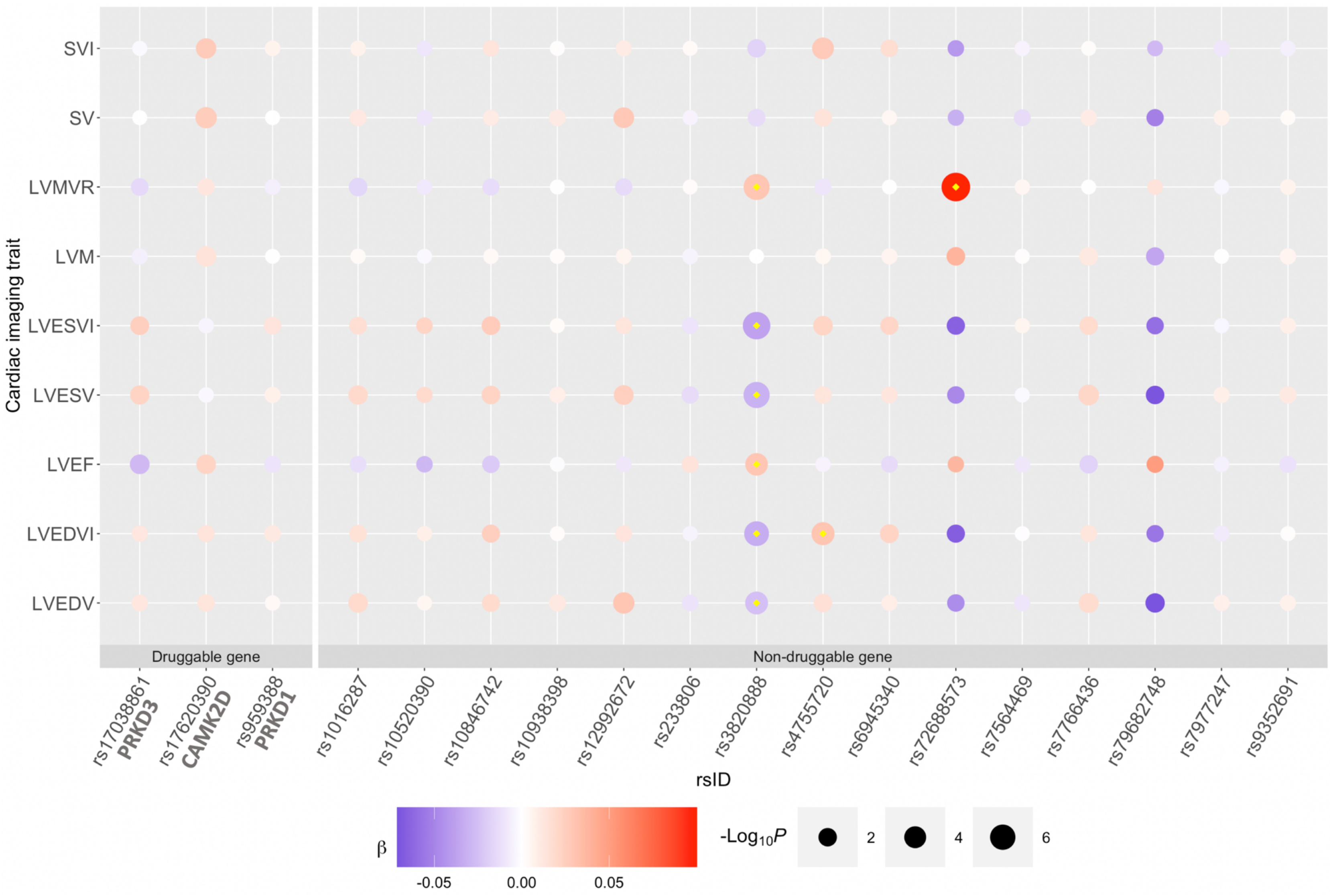


**Supplementary Figure 4. Genetic associations of 18 HF loci against cardiac MRI traits.** The color of the bubble corresponds to the beta coefficient of the genetic association between the loci (x-axis) and trait (y-axis). Blue corresponds to a negative and red corresponds to a positive beta coefficient. The size of each bubble corresponds to the negative logarithm of the association p-value; larger size corresponds to lower p-values. Loci are grouped by druggable and non-druggable genes. Associations which passed the p-value threshold ($p < 1 \times 10^{-4}$ ) are denoted by a yellow diamond. LVEF = Left ventricular ejection fraction, LVEDV = Left ventricular end-diastolic volume, LVEDVI = Left ventricular end-diastolic volume BSA-indexed, LVESV = Left ventricular end-systolic volume, LVESVI = Left ventricular end-systolic volume BSA-indexed, SV = Stroke volume, SVI = Stroke volume BSA-indexed, LVM = left ventricular mass, LVMVR = left ventricular mass to-end-diastolic volume ratio. All tests were two-sided without adjustment for multiple comparisons.

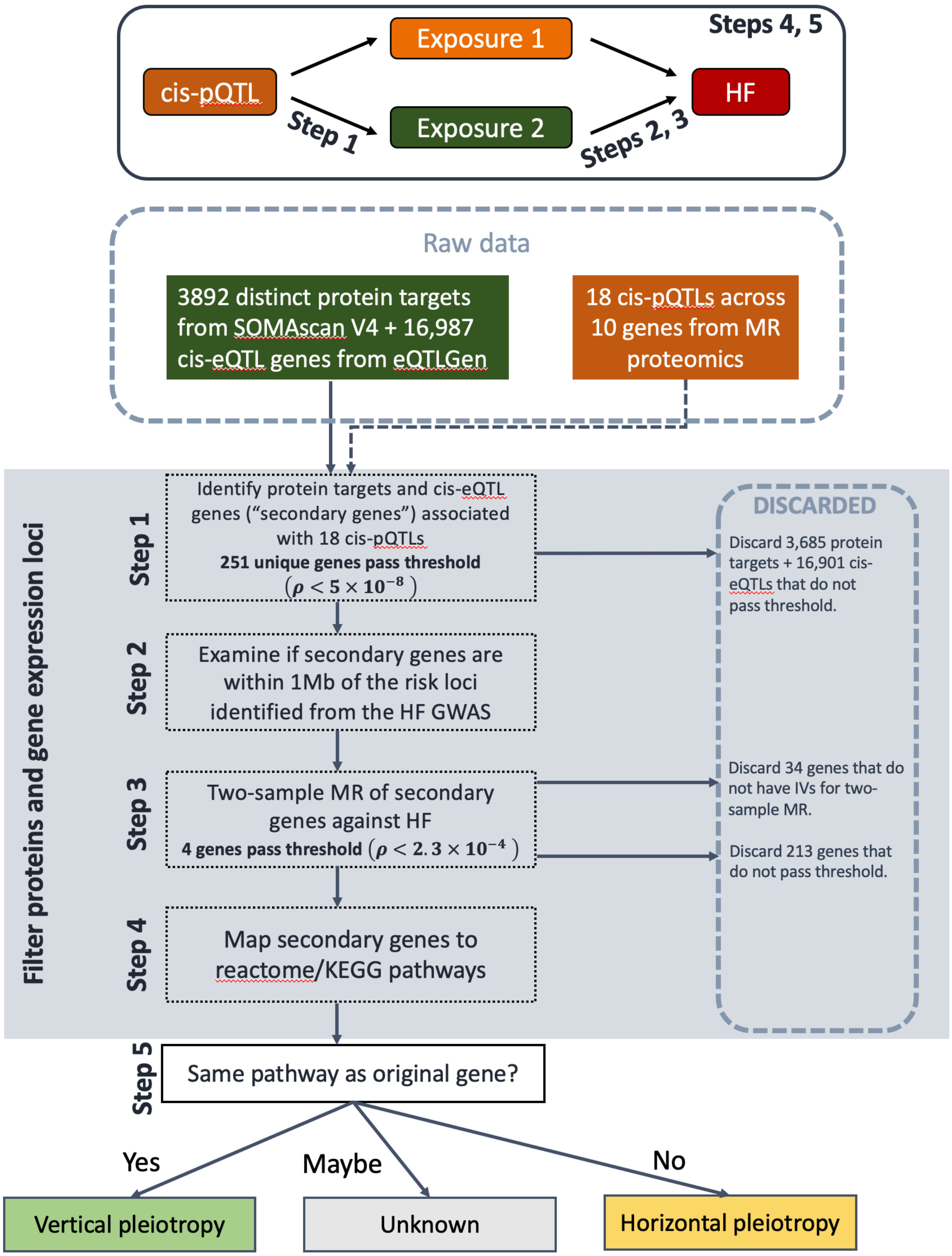


**Supplementary Figure 5. Pipeline of analyses for assessment of horizontal pleiotropy.** For the 18 cis-pQTLs across 10 genes identified from MR-Proteomics for HF, we identified protein targets and cis-eQTL genes ("secondary genes") associated with the 18 cis-pQTLs from 3892 distinct protein targets from SOMAscan V4 and 16,987 cis-eQTL genes derived from eQTLGen (Step 1). We then examined if these secondary genes were within a 1Mb region of risk loci identified from the HF GWAS (Step 2), performed two-sample Mendelian randomization using each as an instrument against HF outcome (Step 3), and mapped genes that passed our significance threshold to reactome/KEGG pathways (Step 4) to determine if the secondary gene was in the same pathway as the original gene— vertical pleiotropy; or not—horizontal pleiotropy (Step 5).

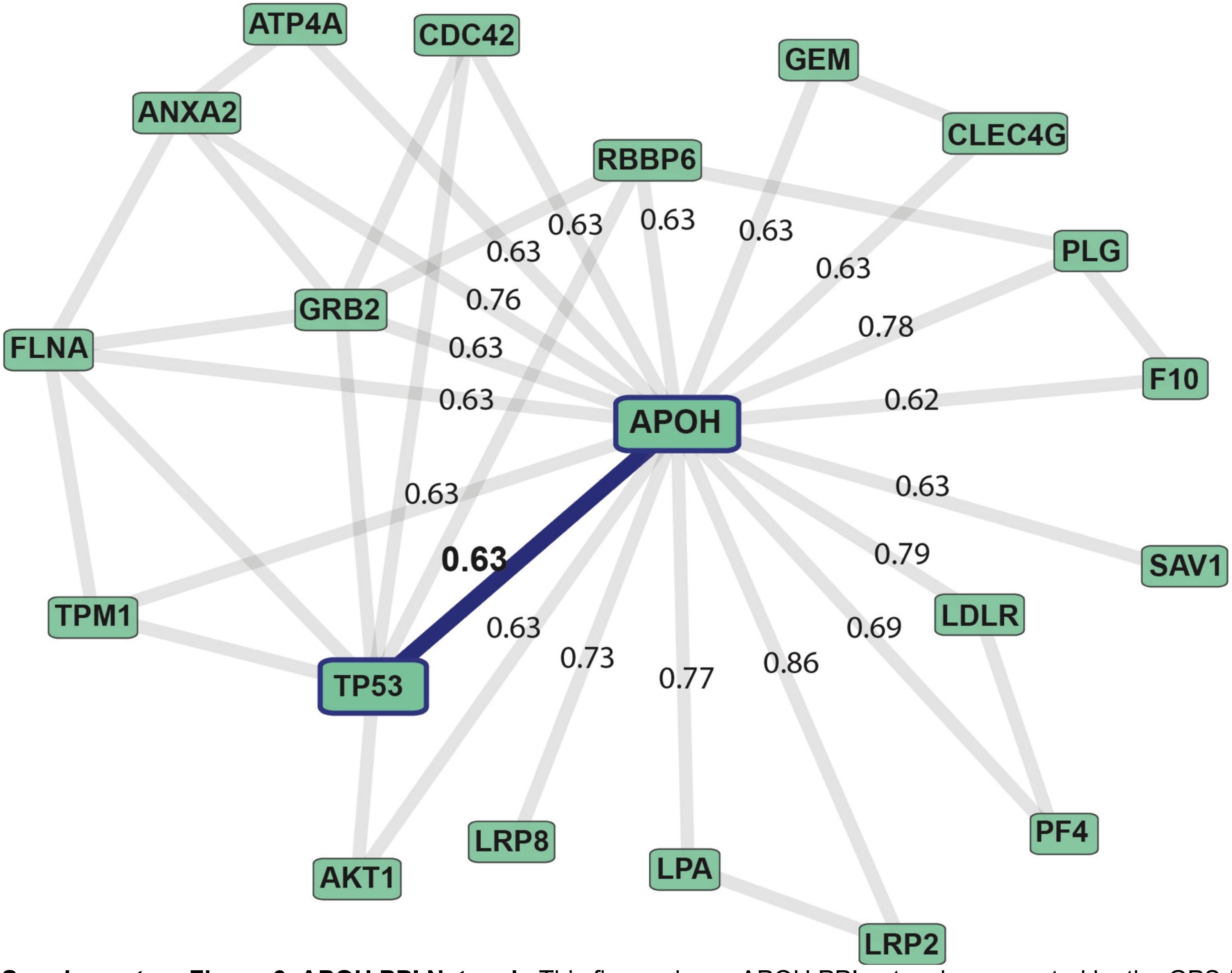


**Supplementary Figure 6. APOH PPI Network.** This figure shows APOH PPI network as reported by the GPS-Prot [PMCID: PMC3213248] data visualization tool (confidence > 0.6). In this database, the APOH-TP53 interaction was extracted from [PMID: 18624398] that is highlighted with a blue edge between APOH and TP53 and edges connected to APOH are annotated by scores from the GPS-Prot APOH analysis.

A

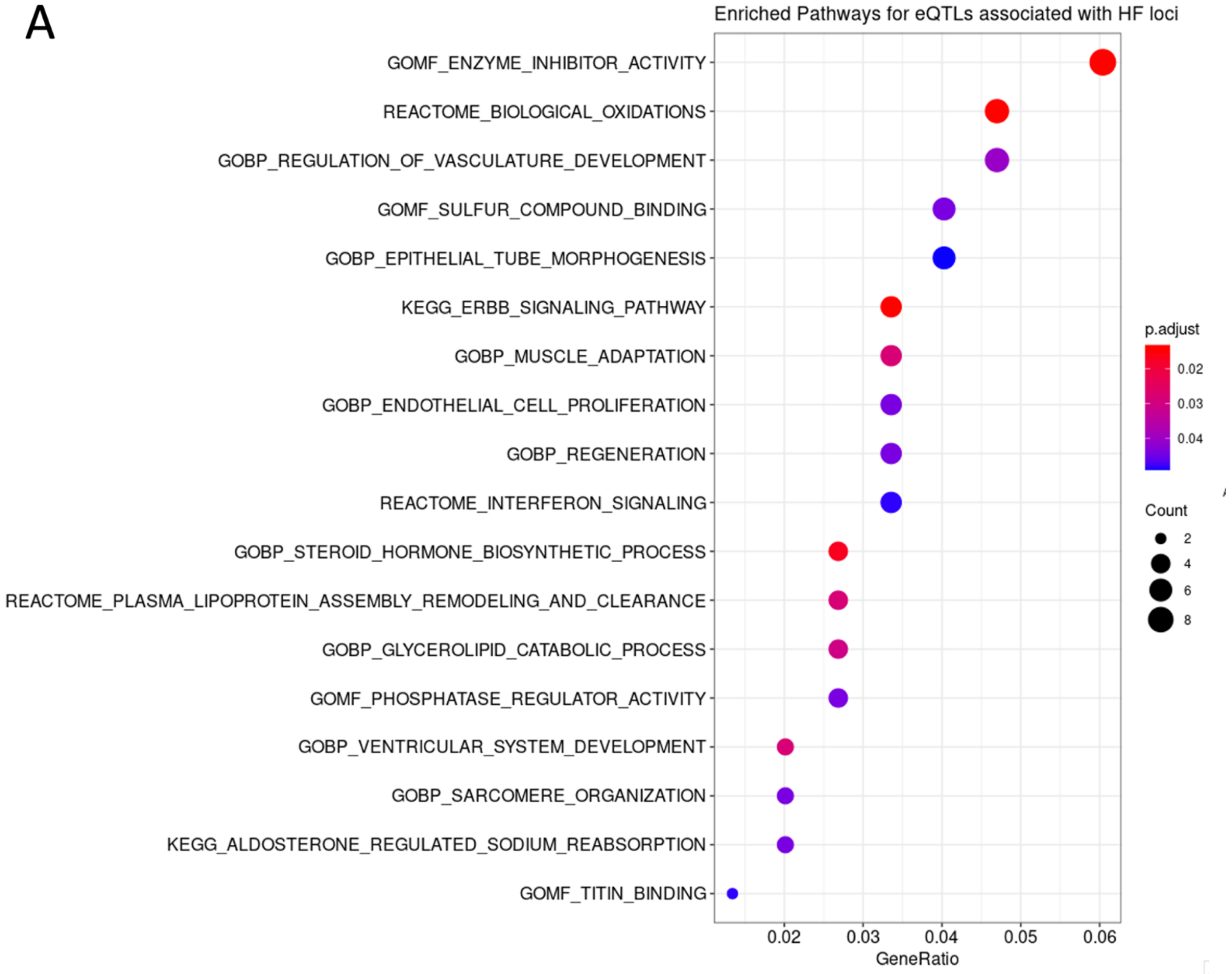


**Supplementary Figure 7. Pathway enrichment analysis of HF variants identified through GWAS and MR proteomics.** Results from over-representation analysis using as input genes for which a genome-wide significant loci or MR instruments is an eQTL. **Panel A**. Selected examples of enriched pathways. All tests were two-sided with adjustment for multiple comparisons; **Panel B (next page)**. Network representation of the communication between observed enriched pathways associated with HF variants.

B

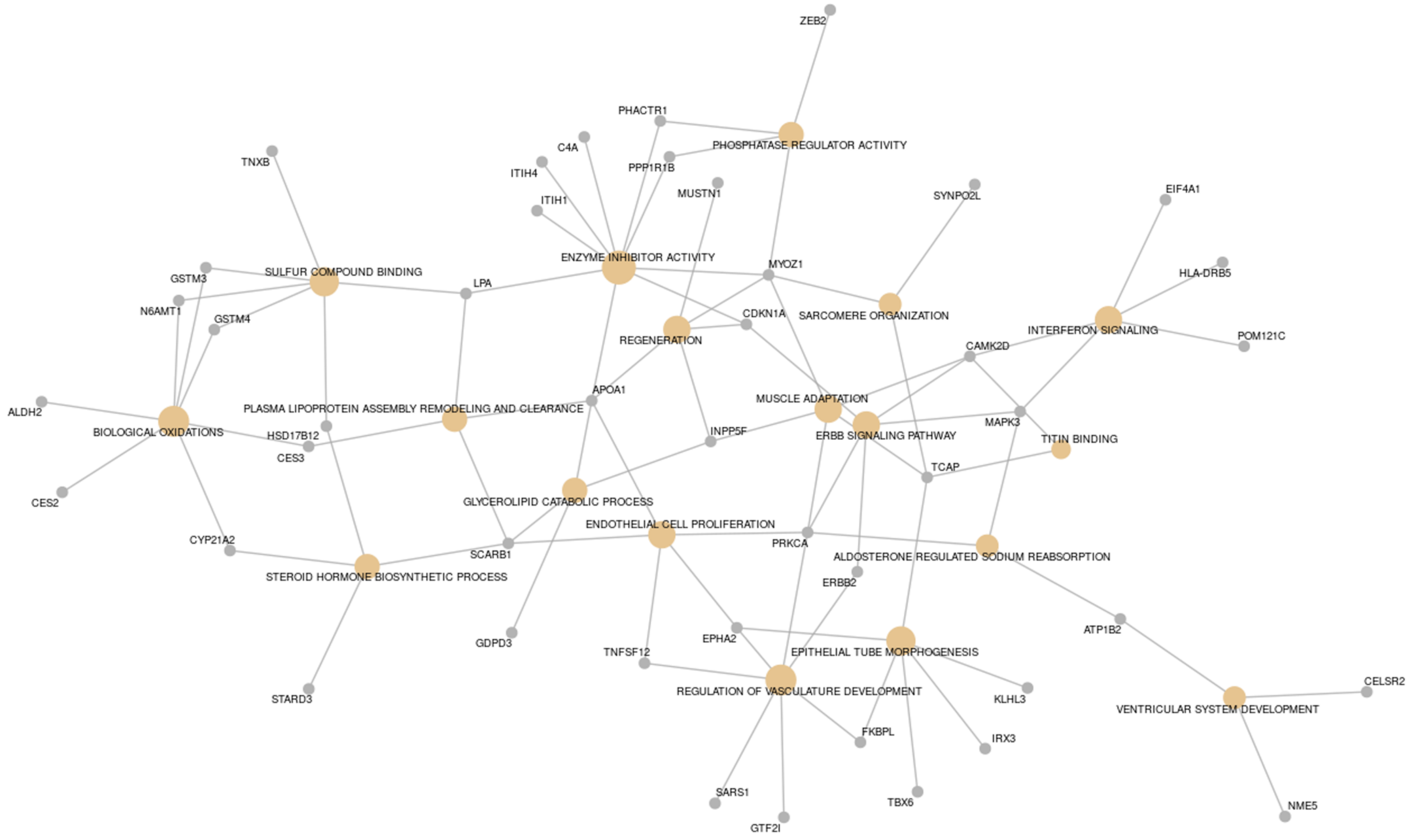
ZEB2
PHACTR1
PHOSPHATASE REGULATOR ACTIVITY
C4A
PPP1R1B
ITIH4
TNXB
MUSTN1
SYNPO2L
EIF4A1
ITIH1
ENZYME INHIBITOR ACTIVITY
MYOZ1
HLA-DRB5
GSTM3
SULFUR COMPOUND BINDING
LPA
N6AMT1
GSTM4
CDKN1A
SARCOMERE ORGANIZATION
INTERFERON SIGNALING
POM121C
REGENERATION
CAMK2D
APOA1
ALDH2
PLASMA LIPOPROTEIN ASSEMBLY REMODELING AND CLEARANCE
MUSCLE ADAPTATION
MAPK3
BIOLOGICAL OXIDATIONS
HSD17B12
INPP5F
ERBB SIGNALING PATHWAY
TITIN BINDING
CES3
TCAP
CES2
GLYCEROLIPID CATABOLIC PROCESS
ENDOTHELIAL CELL PROLIFERATION
CYP21A2
PRKCA
SCARB1
ALDOSTERONE REGULATED SODIUM REABSORPTION
STEROID HORMONE BIOSYNTHETIC PROCESS
ERBB2
ATP1B2
GDPD3
EPHA2
TNFSF12
EPITHELIAL TUBE MORPHOGENESIS
CELSR2
REGULATION OF VASCULATURE DEVELOPMENT
KLHL3
STARD3
VENTRICULAR SYSTEM DEVELOPMENT
FKBPL
IRX3
SARS1
TBX6
NME5
GTF2I

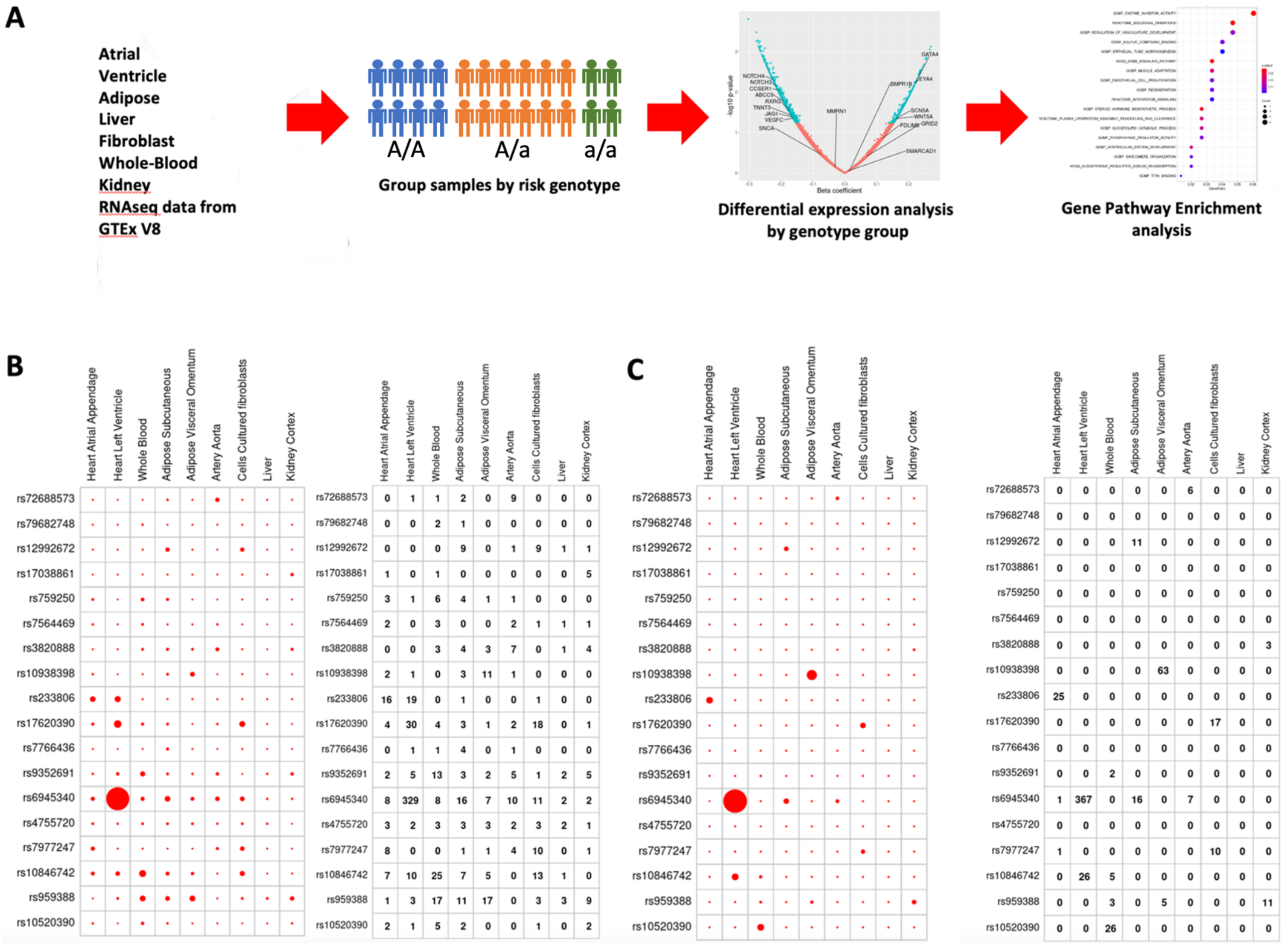


Panel B (number of DE genes):

| | Heart Atrial Appendage | Heart Left Ventricle | Whole Blood | Adipose Subcutaneous | Adipose Visceral Omentum | Artery Aorta | Cells Cultured fibroblasts | Liver | Kidney Cortex |
|---|---|---|---|---|---|---|---|---|---|
| rs72688573 | 0 | 1 | 1 | 2 | 0 | 9 | 0 | 0 | 0 |
| rs79682748 | 0 | 0 | 2 | 1 | 0 | 0 | 0 | 0 | 0 |
| rs12992672 | 0 | 0 | 0 | 9 | 0 | 1 | 9 | 1 | 1 |
| rs17038861 | 1 | 0 | 1 | 0 | 0 | 0 | 0 | 0 | 5 |
| rs759250 | 3 | 1 | 6 | 4 | 1 | 1 | 0 | 0 | 0 |
| rs7564469 | 2 | 0 | 3 | 0 | 0 | 2 | 1 | 1 | 1 |
| rs3820888 | 0 | 0 | 3 | 4 | 3 | 7 | 0 | 1 | 4 |
| rs10938398 | 2 | 1 | 0 | 3 | 11 | 1 | 0 | 0 | 0 |
| rs233806 | 16 | 19 | 0 | 1 | 0 | 0 | 1 | 0 | 0 |
| rs17620390 | 4 | 30 | 4 | 3 | 1 | 2 | 18 | 0 | 1 |
| rs7766436 | 0 | 1 | 1 | 4 | 0 | 1 | 0 | 0 | 0 |
| rs9352691 | 2 | 5 | 13 | 3 | 2 | 5 | 1 | 2 | 5 |
| rs6945340 | 8 | 329 | 8 | 16 | 7 | 10 | 11 | 2 | 2 |
| rs4755720 | 3 | 2 | 3 | 3 | 3 | 2 | 3 | 2 | 1 |
| rs7977247 | 8 | 0 | 0 | 1 | 1 | 4 | 10 | 0 | 1 |
| rs10846742 | 7 | 10 | 25 | 7 | 5 | 0 | 13 | 1 | 0 |
| rs959388 | 1 | 3 | 17 | 11 | 17 | 0 | 3 | 3 | 9 |
| rs10520390 | 2 | 1 | 5 | 2 | 0 | 0 | 1 | 0 | 2 |

Panel C:

| | Heart Atrial Appendage | Heart Left Ventricle | Whole Blood | Adipose Subcutaneous | Adipose Visceral Omentum | Artery Aorta | Cells Cultured fibroblasts | Liver | Kidney Cortex |
|---|---|---|---|---|---|---|---|---|---|
| rs72688573 | 0 | 0 | 0 | 0 | 0 | 6 | 0 | 0 | 0 |
| rs79682748 | 0 | 0 | 0 | 0 | 0 | 0 | 0 | 0 | 0 |
| rs12992672 | 0 | 0 | 0 | 11 | 0 | 0 | 0 | 0 | 0 |
| rs17038861 | 0 | 0 | 0 | 0 | 0 | 0 | 0 | 0 | 0 |
| rs759250 | 0 | 0 | 0 | 0 | 0 | 0 | 0 | 0 | 0 |
| rs7564469 | 0 | 0 | 0 | 0 | 0 | 0 | 0 | 0 | 0 |
| rs3820888 | 0 | 0 | 0 | 0 | 0 | 0 | 0 | 0 | 3 |
| rs10938398 | 0 | 0 | 0 | 0 | 63 | 0 | 0 | 0 | 0 |
| rs233806 | 25 | 0 | 0 | 0 | 0 | 0 | 0 | 0 | 0 |
| rs17620390 | 0 | 0 | 0 | 0 | 0 | 0 | 17 | 0 | 0 |
| rs7766436 | 0 | 0 | 0 | 0 | 0 | 0 | 0 | 0 | 0 |
| rs9352691 | 0 | 0 | 2 | 0 | 0 | 0 | 0 | 0 | 0 |
| rs6945340 | 1 | 367 | 0 | 16 | 0 | 7 | 0 | 0 | 0 |
| rs4755720 | 0 | 0 | 0 | 0 | 0 | 0 | 0 | 0 | 0 |
| rs7977247 | 1 | 0 | 0 | 0 | 0 | 0 | 10 | 0 | 0 |
| rs10846742 | 0 | 26 | 5 | 0 | 0 | 0 | 0 | 0 | 0 |
| rs959388 | 0 | 0 | 3 | 0 | 5 | 0 | 0 | 0 | 11 |
| rs10520390 | 0 | 0 | 26 | 0 | 0 | 0 | 0 | 0 | 0 |

**Supplementary Figure 8. Downstream enrichment analysis of 18 new HF GWAS variants.** Enrichment analysis of differently expressed (DE) genes associated with each of the 18 new HF GWAs loci in different GTEx V8 tissues. **Panel A**. Overall approach. For each selected novel locus we have investigated the downstream transcriptional consequences associated with different genotypes in the following GTEx V8 tissues: heart ventricle, heart atria, adipose omentum, adipose subcutaneous, liver, kidney, whole blood, cultured fibroblasts, aorta. For each tissue we have selected the set of DE genes and conducted over-representation analysis using gene pathways described in GO, KEGG and Reactome. Results were analyzed for each tissue and for each new HF genetic locus. **Panel B**. Number of DE genes per tissue, per variant. All tests were two-sided with adjustment for multiple comparisons**. Panel C**. Number of pathways significantly enriched in each tissue and per loci. Selected pathways needed to have an FDR correct p-value < 0.05 and at least 2 DE genes per tissue. All tests were two-sided with adjustment for multiple comparisons.

**VA Million Veteran Program**
**Core Acknowledgement for Publications**
**Updated December 2021**

**MVP Program Office**

- Program Director - Sumitra Muralidhar, Ph.D.
  US Department of Veterans Affairs, 810 Vermont Avenue NW, Washington, DC 20420
- Associate Director, Scientific Programs - Jennifer Moser, Ph.D.
  US Department of Veterans Affairs, 810 Vermont Avenue NW, Washington, DC 20420
- Associate Director, Cohort Management & Public Relations - Jennifer E. Deen, B.S.
  US Department of Veterans Affairs, 810 Vermont Avenue NW, Washington, DC 20420

**MVP Executive Committee**

- Co-Chair: J. Michael Gaziano, M.D., M.P.H.
  VA Boston Healthcare System, 150 S. Huntington Avenue, Boston, MA 02130
- Co-Chair: Sumitra Muralidhar, Ph.D.
  US Department of Veterans Affairs, 810 Vermont Avenue NW, Washington, DC 20420
- Jean Beckham, Ph.D.
  Durham VA Medical Center, 508 Fulton Street, Durham, NC 27705
- Kyong-Mi Chang, M.D.
  Philadelphia VA Medical Center, 3900 Woodland Avenue, Philadelphia, PA 19104
- Philip S. Tsao, Ph.D.
  VA Palo Alto Health Care System, 3801 Miranda Avenue, Palo Alto, CA 94304
- Shiuh-Wen Luoh, M.D., Ph.D.
  VA Portland Health Care System, 3710 SW US Veterans Hospital Rd, Portland, OR 97239
  US Department of Veterans Affairs, 810 Vermont Avenue NW, Washington, DC 20420
- Juan P. Casas, M.D., Ph.D., Ex-Officio
  VA Boston Healthcare System, 150 S. Huntington Avenue, Boston, MA 02130

**MVP Principal Investigators**

- J. Michael Gaziano, M.D., M.P.H.
  VA Boston Healthcare System, 150 S. Huntington Avenue, Boston, MA 02130
- Philip S. Tsao, Ph.D.
  VA Palo Alto Health Care System, 3801 Miranda Avenue, Palo Alto, CA 94304

**MVP Operations**

- MVP Executive Director – Juan P. Casas, M.D., Ph.D.
  VA Boston Healthcare System, 150 S. Huntington Avenue, Boston, MA 02130
- Director of Regulatory Affairs – Lori Churby, B.S.
  VA Palo Alto Health Care System, 3801 Miranda Avenue, Palo Alto, CA 94304
- MVP Cohort Management Director – Stacey B. Whitbourne, Ph.D.
  VA Boston Healthcare System, 150 S. Huntington Avenue, Boston, MA 02130
- MVP Recruitment/Enrollment Director - Jessica V. Brewer, M.P.H.

VA Boston Healthcare System, 150 S. Huntington Avenue, Boston, MA 02130

- Director, VA Central Biorepository, Boston – Mary T. Brophy M.D., M.P.H.
  VA Boston Healthcare System, 150 S. Huntington Avenue, Boston, MA 02130
- Executive Director for MVP Biorepositories - Luis E. Selva, Ph.D.
  VA Boston Healthcare System, 150 S. Huntington Avenue, Boston, MA 02130
- MVP Informatics, Boston – Shahpoor (Alex) Shayan, M.S.
  VA Boston Healthcare System, 150 S. Huntington Avenue, Boston, MA 02130
- Director, MVP Data Operations/Analytics, Boston – Kelly Cho, M.P.H., Ph.D.
  VA Boston Healthcare System, 150 S. Huntington Avenue, Boston, MA 02130
- Director, Center for Computational and Data Science (C-DACS) & Genomics Core – Saiju Pyarajan Ph.D.
  VA Boston Healthcare System, 150 S. Huntington Avenue, Boston, MA 02130
- Director, Molecular Data Core – Philip S. Tsao, Ph.D.
  VA Palo Alto Health Care System, 3801 Miranda Avenue, Palo Alto, CA 94304
- Director, Phenomics Data Core – Kelly Cho, M.P.H, Ph.D.
  VA Boston Healthcare System, 150 S. Huntington Avenue, Boston, MA 02130
- Director, VA Informatics and Computing Infrastructure (VINCI) – Scott L. DuVall, Ph.D.
  VA Salt Lake City Health Care System, 500 Foothill Drive, Salt Lake City, UT 84148
- MVP Coordinating Centers
  - Cooperative Studies Program Clinical Research Pharmacy Coordinating Center, Albuquerque – Todd Connor, Pharm.D.; Dean P. Argyres, B.S., M.S.
    New Mexico VA Health Care System, 1501 San Pedro Drive SE, Albuquerque, NM 87108
  - Genomics Coordinating Center, Palo Alto – Philip S. Tsao, Ph.D.
    VA Palo Alto Health Care System, 3801 Miranda Avenue, Palo Alto, CA 94304
  - MVP Boston Coordinating Center, Boston - J. Michael Gaziano, M.D., M.P.H.
    VA Boston Healthcare System, 150 S. Huntington Avenue, Boston, MA 02130
  - MVP Information Center, Canandaigua – Brady Stephens, M.S.
    Canandaigua VA Medical Center, 400 Fort Hill Avenue, Canandaigua, NY 14424

**Current MVP Local Site Investigators**

- Atlanta VA Medical Center (Peter Wilson, M.D.)
  1670 Clairmont Road, Decatur, GA 30033
- Bay Pines VA Healthcare System (Rachel McArdle, Ph.D.)
  10,000 Bay Pines Blvd Bay Pines, FL 33744
- Birmingham VA Medical Center (Louis Dellitalia, M.D.)
  700 S. 19th Street, Birmingham AL 35233
- Central Western Massachusetts Healthcare System (Kristin Mattocks, Ph.D., M.P.H.)
  421 North Main Street, Leeds, MA 01053
- Cincinnati VA Medical Center (John Harley, M.D., Ph.D.)
  3200 Vine Street, Cincinnati, OH 45220

- Clement J. Zablocki VA Medical Center (Jeffrey Whittle, M.D., M.P.H.)
  5000 West National Avenue, Milwaukee, WI 53295
- VA Northeast Ohio Healthcare System (Frank Jacono, M.D.)
  10701 East Boulevard, Cleveland, OH 44106
- Durham VA Medical Center (Jean Beckham, Ph.D.)
  508 Fulton Street, Durham, NC 27705
- Edith Nourse Rogers Memorial Veterans Hospital (John Wells., Ph.D.)
  200 Springs Road, Bedford, MA 01730
- Edward Hines, Jr. VA Medical Center (Salvador Gutierrez, M.D.)
  5000 South 5th Avenue, Hines, IL 60141
- Veterans Health Care System of the Ozarks (Kathrina Alexander, M.D.)
  1100 North College Avenue, Fayetteville, AR 72703
- Fargo VA Health Care System (Kimberly Hammer, Ph.D.)
  2101 N. Elm, Fargo, ND 58102
- VA Health Care Upstate New York (James Norton, Ph.D.)
  113 Holland Avenue, Albany, NY 12208
- New Mexico VA Health Care System (Gerardo Villareal, M.D.)
  1501 San Pedro Drive, S.E. Albuquerque, NM 87108
- VA Boston Healthcare System (Scott Kinlay, M.B.B.S., Ph.D.)
  150 S. Huntington Avenue, Boston, MA 02130
- VA Western New York Healthcare System (Junzhe Xu, M.D.)
  3495 Bailey Avenue, Buffalo, NY 14215-1199
- Ralph H. Johnson VA Medical Center (Mark Hamner, M.D.)
  109 Bee Street, Mental Health Research, Charleston, SC 29401
- Columbia VA Health Care System (Roy Mathew, M.D.)
  6439 Garners Ferry Road, Columbia, SC 29209
- VA North Texas Health Care System (Sujata Bhushan, M.D.)
  4500 S. Lancaster Road, Dallas, TX 75216
- Hampton VA Medical Center (Pran Iruvanti, D.O., Ph.D.)
  100 Emancipation Drive, Hampton, VA 23667
- Richmond VA Medical Center (Michael Godschalk, M.D.)
  1201 Broad Rock Blvd., Richmond, VA 23249
- Iowa City VA Health Care System (Zuhair Ballas, M.D.)
  601 Highway 6 West, Iowa City, IA 52246-2208
- Eastern Oklahoma VA Health Care System (River Smith, Ph.D.)
  1011 Honor Heights Drive, Muskogee, OK 74401
- James A. Haley Veterans' Hospital (Stephen Mastorides, M.D.)
  13000 Bruce B. Downs Blvd, Tampa, FL 33612
- James H. Quillen VA Medical Center (Jonathan Moorman, M.D., Ph.D.)
  Corner of Lamont & Veterans Way, Mountain Home, TN 37684
- John D. Dingell VA Medical Center (Saib Gappy, M.D.)
  4646 John R Street, Detroit, MI 48201
- Louisville VA Medical Center (Jon Klein, M.D., Ph.D.)

800 Zorn Avenue, Louisville, KY 40206

- Manchester VA Medical Center (Nora Ratcliffe, M.D.)
  718 Smyth Road, Manchester, NH 03104
- Miami VA Health Care System (Ana Palacio, M.D., M.P.H.)
  1201 NW 16th Street, 11 GRC, Miami FL 33125
- Michael E. DeBakey VA Medical Center (Olaoluwa Okusaga, M.D.)
  2002 Holcombe Blvd, Houston, TX 77030
- Minneapolis VA Health Care System (Maureen Murdoch, M.D., M.P.H.)
  One Veterans Drive, Minneapolis, MN 55417
- N. FL/S. GA Veterans Health System (Peruvemba Sriram, M.D.)
  1601 SW Archer Road, Gainesville, FL 32608
- Northport VA Medical Center (Shing Shing Yeh, Ph.D., M.D.)
  79 Middleville Road, Northport, NY 11768
- Overton Brooks VA Medical Center (Neeraj Tandon, M.D.)
  510 East Stoner Ave, Shreveport, LA 71101
- Philadelphia VA Medical Center (Darshana Jhala, M.D.)
  3900 Woodland Avenue, Philadelphia, PA 19104
- Phoenix VA Health Care System (Samuel Aguayo, M.D.)
  650 E. Indian School Road, Phoenix, AZ 85012
- Portland VA Medical Center (David Cohen, M.D.)
  3710 SW U.S. Veterans Hospital Road, Portland, OR 97239
- Providence VA Medical Center (Satish Sharma, M.D.)
  830 Chalkstone Avenue, Providence, RI 02908
- Richard Roudebush VA Medical Center (Suthat Liangpunsakul, M.D., M.P.H.)
  1481 West 10th Street, Indianapolis, IN 46202
- Salem VA Medical Center (Kris Ann Oursler, M.D.)
  1970 Roanoke Blvd, Salem, VA 24153
- San Francisco VA Health Care System (Mary Whooley, M.D.)
  4150 Clement Street, San Francisco, CA 94121
- South Texas Veterans Health Care System (Sunil Ahuja, M.D.)
  7400 Merton Minter Boulevard, San Antonio, TX 78229
- Southeast Louisiana Veterans Health Care System (Joseph Constans, Ph.D.)
  2400 Canal Street, New Orleans, LA 70119
- Southern Arizona VA Health Care System (Paul Meyer, M.D., Ph.D.)
  3601 S 6th Avenue, Tucson, AZ 85723
- Sioux Falls VA Health Care System (Jennifer Greco, M.D.)
  2501 W 22nd Street, Sioux Falls, SD 57105
- St. Louis VA Health Care System (Michael Rauchman, M.D.)
  915 North Grand Blvd, St. Louis, MO 63106
- Syracuse VA Medical Center (Richard Servatius, Ph.D.)
  800 Irving Avenue, Syracuse, NY 13210
- VA Eastern Kansas Health Care System (Melinda Gaddy, Ph.D.)
  4101 S 4th Street Trafficway, Leavenworth, KS 66048

- VA Greater Los Angeles Health Care System (Agnes Wallbom, M.D., M.S.)
  11301 Wilshire Blvd, Los Angeles, CA 90073
- VA Long Beach Healthcare System (Timothy Morgan, M.D.)
  5901 East 7th Street Long Beach, CA 90822
- VA Maine Healthcare System (Todd Stapley, D.O.)
  1 VA Center, Augusta, ME 04330
- VA New York Harbor Healthcare System (Peter Liang, M.D., M.P.H.)
  423 East 23rd Street, New York, NY 10010
- VA Pacific Islands Health Care System (Daryl Fujii, Ph.D.)
  459 Patterson Rd, Honolulu, HI 96819
- VA Palo Alto Health Care System (Philip Tsao, Ph.D.)
  3801 Miranda Avenue, Palo Alto, CA 94304-1290
- VA Pittsburgh Health Care System (Patrick Strollo, Jr., M.D.)
  University Drive, Pittsburgh, PA 15240
- VA Puget Sound Health Care System (Edward Boyko, M.D.)
  1660 S. Columbian Way, Seattle, WA 98108-1597
- VA Salt Lake City Health Care System (Jessica Walsh, M.D.)
  500 Foothill Drive, Salt Lake City, UT 84148
- VA San Diego Healthcare System (Samir Gupta, M.D., M.S.C.S.)
  3350 La Jolla Village Drive, San Diego, CA 92161
- VA Sierra Nevada Health Care System (Mostaqul Huq, Pharm.D., Ph.D.)
  975 Kirman Avenue, Reno, NV 89502
- VA Southern Nevada Healthcare System (Joseph Fayad, M.D.)
  6900 North Pecos Road, North Las Vegas, NV 89086
- VA Tennessee Valley Healthcare System (Adriana Hung, M.D., M.P.H.)
  1310 24th Avenue, South Nashville, TN 37212
- Washington DC VA Medical Center (Jack Lichy, M.D., Ph.D.)
  50 Irving St, Washington, D. C. 20422
- W.G. (Bill) Hefner VA Medical Center (Robin Hurley, M.D.)
  1601 Brenner Ave, Salisbury, NC 28144
- White River Junction VA Medical Center (Brooks Robey, M.D.)
  163 Veterans Drive, White River Junction, VT 05009
- William S. Middleton Memorial Veterans Hospital (Prakash Balasubramanian, M.D.)
  2500 Overlook Terrace, Madison, WI 53705
